# PGT-A Mosaicism Reporting Lacks Clinical Predictive Value: Evidence from a Multisite, Double-Blinded Study with Independent Validation Across 15,315 Single-Embryo Transfers

**DOI:** 10.1101/2025.06.23.25329758

**Authors:** P. Gill, Xin Tao, Yiping Zhan, F. Mulas, C.S. Ottolini, L. Picchetta, S. Caroselli, D. Babariya, Dagan Wells, Georgina Clark, Elena Fernandez Marcos, Carlos Marin Vallejo, Vaidehi Jobanputra, Marie Werner, Richard Scott, Thomas Molinaro, Josep Pla, Vanessa Vergara Bravo, Antonio Requena Miranda, Juan Antonio García Velasco, Antonio Pellicer, Emily Mounts, Chaim Jalas, Antonio Capalbo

**Affiliations:** IVIRMA, Clinical Research, Basking Ridge, New Jersey; Juno Genetics, Genetic Lab, Basking Ridge, New Jersey, US; Juno Genetics, Reproductive Genetics, Rome, Italy; University College London-Institute for Women’s Health, Department of Maternal and Fetal Medicine, London, United Kingdom; University of Teramo, Department of Department of Bioscience and Agro-Food and Environmental Technology.; Juno Genetics, Hayakawa Building, Oxford Science Park, Oxford, OX4 4GB, UK; University of Oxford, Nuffield Department of Women’s and Reproductive Health, John Radcliffe Hospital, Oxford, OX3 9DU, UK; Juno Genetics, Genetic Lab, Valencia, Spain; Department of Pathology and Cell Biology, Columbia University Irving Medical Center, New York, NY 10032; IVIRMA Global Research Alliance, New Jersey, Basking Ridge, NJ, USA; Foundation for Embryonic Competence, Basking Ridge, New Jersey; Division of Reproductive Endocrinology, Department Obstetrics and Gynecology, Yale University School of Medicine, New Haven, Connecticut; Reproductive Genetics Unit, IVI-RMA Global, Barcelona, Spain; IVRMA Global Research Alliance Madrid, Madrid, Spain; IVIRMA Global Research Alliance, IVIRMA Rome, Roma, Italy; Unit of Molecular Genetics, Center for Advanced Studies and Technology (CAST), “G. D’Annunzio” University of Chieti-Pescara, Chieti, Italy; IVI Foundation, Health Research Institute La Fe, Valencia, Spain

## Abstract

Preimplantation genetic testing for aneuploidy (PGT-A) aims to improve IVF outcomes by identifying embryos with lethal chromosomal abnormalities leading to impaired embryonic development and pregnancy loss, particularly with advancing maternal age. The adoption of next-generation sequencing (NGS) has enabled the detection of subtler chromosomal variations of uncertain clinical significance, such as intermediate copy number (ICN), leading to the classification of putative mosaicism.

To assess the clinical utility of mosaicism reporting in PGT, we conducted a large, multi-site, double-blinded, non-selection study across U.S. fertility clinics (Feb 2020–Oct 2022), analyzing 9,828 single embryo transfers (SETs) from 7,564 IVF cycles. The findings were independently validated using a separate dataset of 5,487 euploid SETs from European clinics (May 2022–March 2024), representing a distinct patient population with different clinical characteristics. This design allowed for ICN values to be unblinded post-transfer, enabling unbiased comparisons between embryos that, under a mosaic reporting framework, would have been labeled as mosaic or euploid. In both cohorts, the presence of ICN showed no significant impact on clinical outcomes when mosaicism status was blinded and evaluated alongside established clinical and embryological parameters. AUROC analysis revealed no meaningful difference in predictive performance between models with and without ICN data (AUC = 0.552 vs. 0.555; 0.569 vs. 0.578; all P > 0.05). Miscarriage, obstetric, and neonatal outcomes were also comparable. These findings strongly suggest that reporting putative mosaicism based on ICN lacks clinical utility and should not be used to inform embryo selection policies.

## Introduction

The prevalence of chromosomal aneuploidies is the primary determinant of an embryo’s ability to develop to term (Capalbo et al., 2022; Gruhn et al., 2019; Tiegs et al., 2021). Aneuploidy, arising during gametogenesis (predominantly maternal but also paternal) (Rana et al., 2023), has long been associated with pregnancy loss, chromosomally abnormal gestations, and, most frequently, failure of embryo implantation (Hassold et al., 1980; Kung et al., 2015). Seminal studies have demonstrated a strong positive correlation between maternally derived aneuploidies and aging, progressively driving the decline in human fertility across the reproductive lifespan (Franasiak et al., 2014; Gruhn et al., 2019).

Given the profound implications of aneuploidy on reproductive success, especially with advancing maternal age, preimplantation genetic testing for aneuploidy (PGT-A) was introduced as an adjunct to in vitro fertilization (IVF) with the premise of identification and deselection of embryos incompatible with normal development (Verlinsky et al., 1990). When correctly validated, routine use of PGT-A has shown high predictive value for negative clinical outcomes associated with uniform whole chromosome aneuploidy detected in clinical trophectoderm (TE) biopsies (Tiegs et al., 2021). In turn, incorporating PGT-A into IVF cycles has been associated with significant clinical benefits, including reduced miscarriage rates (Mumusoglu et al., 2025) and a lower risk of multiple pregnancies (Ubaldi et al., 2015), while maintaining similar IVF success rates per cycle started (Sacchi et al., 2019). As a result, PGT-A has seen increased adoption in IVF practices worldwide, now utilized in approximately 50% of IVF cycles in the US (Hipp et al., 2022).

A combination of contemporary PGT-A workflows, involving biopsy and analysis of multi-cell samples from the TE of developing blastocyst-stage embryos and significant improvements in DNA quantitative analysis, especially through next-generation sequencing (NGS), have facilitated the potential detection of more subtle chromosomal variations, leading to the interpretation of chromosomal mosaicism (Munné et al., 2017; Treff & Marin, 2021). Chromosomal mosaicism, which differs from aneuploidy arising in gametogenesis, is traditionally defined by the presence of at least two karyotypically distinct cell lines within the embryo. In PGT-A, mosaicism is commonly inferred when the chromosome copy number falls outside the normal disomic range but does not reach thresholds for monosomy or trisomy (Capalbo et al., 2017; Treff & Marin, 2021). The intermediate copy number (ICN) observed in such cases suggests deviation from the disomy state, but that not all cells in the biopsy are aneuploid (Capalbo et al., 2021).

The goal of PGT-A has classically been to identify uniform gross chromosomal aneuploidies largely incompatibly with life and exclude such embryos from the pool available for transfer. In turn – assuming minimal risks of the associated embryology biopsy and workflow and a low false-positive rate of PGT-A - the process would be expected to improve the efficiency of IVF. However, the routine clinical reporting of putative mosaic findings in PGT-A – namely, through the use of arbitrarily selected cutoffs to define “mosaic” result by ICNs (vs “euploid” and “aneuploid”) categories – has caused major confusion about best practices for ranking embryos for transfer. Arbitrary thresholds for ICN and lack of properly designed analytical and clinical validation studies have led to classification inconsistencies across laboratories, with embryos called “euploid” in one lab potentially being classified as “mosaic” in another, leading to vastly different rates of mosaicism reporting across PGT laboratories (2-20%) (Bardos et al., 2023; Girardi et al., 2023; Goodrich et al., 2016; Popovic et al., 2024). Additionally, the lack of reproducibility of mosaicism detection when embryos undergo a second biopsy suggests that the ICN detected in clinical TE biopsy in the vast majority of cases does not reflect the chromosomal status of the entire embryo (Capalbo et al., 2017; J. Kim et al., 2022) and in part may be an artifact of certain PGT-A platforms, leading to misclassification of both euploid and aneuploid embryos as mosaic (Capalbo et al., 2021; Popa et al., 2025).

In recent years there have been numerous reports of healthy term deliveries following the transfer of putative mosaic embryos (Greco et al., 2015; Viotti et al., 2023). Despite this, or perhaps because of it, vigorous debate continues regarding the clinical relevance of embryos with instances of putative mosaicism detected by PGT-A. While the ASRM Practice Committee has issued an opinion on the clinical management of putative mosaic PGT-A results (“Clinical Management of Mosaic Results from Preimplantation Genetic Testing for Aneuploidy of Blastocysts: A Committee Opinion,” 2023), there is no professional consensus on the optimal testing, reporting, or prioritization strategy. Although there has been increasing acceptance of mosaic embryo transfer in the U.S. (“Clinical Management of Mosaic Results from Preimplantation Genetic Testing for Aneuploidy of Blastocysts: A Committee Opinion,” 2023; T. Kim et al., 2018; Viotti et al., 2023), there is evidence that embryos reported as mosaic are frequently disallowed for transfer by IVF clinicians and are therefore discarded, resulting in loss of potential healthy live births (ESHRE Working Group on Chromosomal Mosaicism et al., 2022).

Two primary study designs can be employed to assess the clinical significance of putative mosaicism detected by PGT-A. The first approach involves retrospective, unblinded studies, which report clinical outcomes following the transfer of embryos already known and classified as mosaic. To date, such studies have generally shown lower live birth rates and higher miscarriage rates associated with the transfer of putative mosaic embryos (Viotti et al., 2023). However, it is important to acknowledge the significant selection bias inherent to this design: because these embryos are not classified as “normal” or euploid, they are typically only considered for transfer after failed transfers of euploid embryos or when no euploid embryos are available. Consequently, mosaic embryo transfers are often performed in a poor-prognosis population, which limits the generalizability of the findings.

In contrast, prospective blinded studies offer a more robust design to assess the significance of ICN, since only results of already proven clinical significance (e.g. whole chromosome aneuploidy) are reported, while mosaic findings are not. In these studies, embryo selection is based solely on morphological criteria, consistent with routine clinical practice, and without any influence from genetic reporting. This design eliminates selection bias and provides a more evidence-based assessment of the true reproductive potential of embryos with ICNs or any other emerging biomarkers. To date, the only published study employing this approach demonstrated that embryos classified as having low- or medium-level mosaicism (defined as <50% abnormal cells in the TE biopsy specimen) showed comparable developmental and reproductive outcomes to euploid embryos, with no evidence of mosaicism detected in postnatal genetic analyses (Capalbo et al., 2021).

In 2021, the clinical validation of the PGTSeq platform in diagnosing whole chromosome aneuploidy was completed through a blinded diagnostic accuracy non-selection study (Tiegs et al., 2021). In turn, a structured clinical reporting strategy was adopted by the laboratory in which only uniform (non-mosaic) aneuploidy is reported as a default. Mosaic findings of any level and on any chromosome are considered secondary findings of uncertain clinical significance and are not reported unless specifically requested by clinics through an “opt-in” on the test requisition. While not the primary goal of this clinical approach, the policy of default reporting of only uniform aneuploidy findings secondarily enabled the prospective generation of a unique dataset of blinded embryo transfer outcomes, in which embryos classified as negative for uniform aneuploidies—some of which are euploid, and others with mosaic findings based on ICN and supporting SNP genotyping data—were all transferred without bias.

The primary objective of this study, therefore, was to assess and compare the clinical outcomes of euploid embryos versus putative mosaic embryos of all types in a prospective, double-blinded, non-selection study design spanning multiple IVF clinics in the United States, and to independently validate the results using a dataset from a European IVF setting. This unique study design enabled the unblinding of ICN values after embryo transfer, allowing for an unbiased comparison of clinical outcomes between those embryos that - in a mosaic reporting scheme - would have been classified as putative mosaic, and those that would have been reported as euploid. Furthermore, the blinded design enabled investigation of the clinical value of reporting putative mosaicism findings in conjunction with established clinical features already understood to influence IVF outcomes. As the largest systematic evaluation of the clinical significance and utility of mosaicism reporting in PGT-A to date, the methodology and size of this data set are intended to provide critical insights to inform best clinical practices of PGT-A in the future.

## Materials and Methods

### Study design

This multi-site, blinded, prospective cohort study was conducted between February 2020 and October 2022 across five participating fertility clinics in the United States (primary cohort). All patients underwent in vitro fertilization (IVF) with intracytoplasmic sperm injection (ICSI), followed by TE biopsy and preimplantation genetic testing for aneuploidy (PGT-A).

The experimental group consisted of patients with at least one embryo transferred that was negative for uniform whole chromosome aneuploidy (WCA) and segmental aneuploidy (SA), in which putative mosaic findings were present (though not reported). The control group consisted of patients with at least one embryo transferred which was negative for uniform WCA and SA and negative for putative mosaic findings (euploid). All patients underwent a single frozen embryo transfer (FET), and post-transfer outcomes were assessed after unblinding the ICN status of the transferred embryos. Institutional Review Board (IRB) approval for this primary phase with the US datasets was obtained to evaluate post-embryo transfer outcomes following unblinding (Advarra IRB, Pro00064197).

To validate and corroborate these findings in a diverse patient population and clinical setting, outcomes of 5,487 single euploid embryo transfer cycles were collected across 17 participating fertility clinics in Spain from May 2022 to March 2024 (validation cohort). In this European validation group, the clinical protocol in these centers was aligned with US clinical practice, reporting only uniform WCA and SA findings, and experimental and control groups were as defined above. Ethical Committee (EC) approval for this validation phase with the European datasets was obtained (CEIM-HOSPITAL UNIVERSITARIO Y POLITECNICO LA FE, 1064-1). Data recording was completed in January 2025 for the European datasets with available obstetrical and neonatal outcomes at that time.

### Participants and IVF procedures

For both the primary and validation cohorts, embryos generated in all IVF cycles from the participating clinics were considered, including those in which an oocyte and/or sperm donor or a gestational carrier were utilized. All oocyte providers were between 18-45 years old at the time of oocyte collection and embryos were transferred to women with a BMI <45 kg/m^2^ at the time of the procedure. Embryos from patients undergoing PGT for monogenic disorders or structural chromosome rearrangements were excluded from the study.

All participants received implications counselling for PGT-A by their care team before undertaking IVF treatment. As part of informed consent, patients were notified that the standard PGT-A clinical approach included reporting of uniform WCA and SA only, and that samples with putative evidence of mosaicism (based on interpretation of ICN and SNP data, as described below) would be reported, together with euploid-result samples, as “negative for aneuploidy”. All patients were counseled on the limitations of PGT-A and encouraged to proceed with routine prenatal screening and testing in pregnancy as per standard clinical practice guidelines.

IVF procedures were carried out according to standard practices employed at each clinic for both the primary and the validation cohorts. Decisions regarding the type of stimulation protocol, dosing of medications, and trigger administration were at the discretion of the patient’s physician and followed standard clinical practice. Oocyte retrieval was performed via ultrasound-guided aspiration approximately 36 hours after triggering final oocyte maturation. ICSI was performed on all mature metaphase II oocytes. Zygotes displaying two pronuclei were then cultured through to the blastocyst stage and PGT-A was performed via TE biopsy prior to cryopreservation. Embryos were biopsied on day 5, 6, or 7 if they reached the blastocyst stage and were at least grade CC or higher based on the modified Gardner scoring system (Gardner & Schoolcraft, 1999). Single FET was performed in a subsequent cycle according to the practice of each center. The blastocysts were evaluated and selected by the embryologist for transfer based on standard morphological criteria, blinded to any putative mosaicism findings. The transfer was performed after obtaining adequate endometrial proliferation (preferably >6mm before initiating progesterone) via a medicated or modified natural cycle protocol. Progesterone supplementation was continued until ten weeks gestational age. The IVF procedures performed at each clinical site are not expected to influence the outcomes of this study, as they were applied equally and independently to both the experimental and control arms.

### PGTSeq-A and mosaicism reporting

All embryo biopsies were sent to one of three testing sites (Juno Genetics US for the US primary cohort and Juno Genetics Spain or UK for the European validation cohort) for analysis and underwent testing using the same PGTSeq-A assay (Tiegs et al., 2021). The PGTSeq-A assay is a custom, targeted NGS platform with ∼5000 loci across the human genome that provides two data outputs: quantitative based on chromosome copy number (log2 ratio plot) and qualitative based on genotyping of single nucleotide polymorphisms (SNPs) (to compute the alternative allele fraction) to inform the uniform aneuploid classification. This assay has undergone extensive preclinical validation on cell lines and embryo rebiopsies (J. Kim et al., 2022), as well as a unique and fundamental clinical validation performed via a prospective, blinded, study showing lack of sustained implantation potential for embryos diagnosed as uniform aneuploid (Tiegs et al., 2021). Nonetheless, to further evaluate the analytical capability and consistency of mosaicism detection in this PGT-A assay, 10% incremental variations in ICN were assessed through cell line mixing experiments (Supplementary Table 1).

Raw PGTSeq-A data were unblinded post-transfer and the copy number value was assessed for each autosome (chromosome 1-22) along with the corresponding SNP profile. Putative mosaicism was called when an ICN did not reach the criteria for a full monosomy (loss) or trisomy (gain) and when there was evidence of supporting allelic ratios in the SNP genotyping analysis (see Supplementary Figure 1 for details). Euploidy was called when the ICN and SNP allelic ratios together did not meet criteria for whole chromosome or segmental mosaicism (negative for ICN). Patients who transferred putative mosaic embryos (positive for ICN) comprised the experimental group, while patients who transferred euploid embryos (negative for ICN) comprised the control group.

In the experimental group - in which results had demonstrated an ICN in one or more autosomes – outcomes were classified according to subcategorizations of putative mosaic findings as follows: (1) “whole chromosome ICN” when at least one whole chromosome was involved (wcICN); (2) “segmental ICN” (sICN) when at least one segment (less than a full chromosome and down to approximately 3-7 Mb in size) of the chromosome(s) was involved; and (3) “complex ICN” when a combination of wcICN and sICN were identified.

For the main analysis ICN was considered as binary variable (i.e., presence or absence of ICN). In the sub-analysis, ICN was analyzed as a continuous variable to identify potential copy number threshold values associated with significant differences in clinical outcomes. Embryos with X or Y chromosome ICNs were excluded from the full data analysis because of the inability of using the mosaicism prediction in isolation for the two chromosomes. However, descriptive statistics were calculated for clinical outcomes and reported in the manuscript.

### Outcome measures

The primary outcome for investigation in the study was live birth rate, defined as delivery after 24 weeks gestational age (Zegers-Hochschild et al., 2017). Secondary outcomes included clinical pregnancy rate (defined as by presence of gestational sac observed by vaginal ultrasound), clinical miscarriage rate (defined as loss of a clinical pregnancy before 24 weeks of gestation, including stillbirths), and adverse pregnancy outcomes (maternal and neonatal). Maternal adverse outcomes analyzed included hypertensive disorders of pregnancy, gestational diabetes and placental abruption. Neonatal adverse outcomes included preterm delivery (delivery before 37 weeks gestational age), low birth weight (<2500g), NICU admission, and congenital anomalies. Adverse outcomes were self-reported by patients who were contacted via telephone and/or email post-delivery by each clinic as per routine protocol to obtain delivery outcomes.

### Statistical analysis

The primary endpoint of this analysis is the equivalence assessment in the live birth rate (LBR) when comparing embryos without ICNs (euploid) to those with ICNs (putative mosaic). To estimate the distribution of euploid and putative mosaic embryos in the study cohort, historical data was referenced; this indicated that approximately 15% of embryos exhibited various ICNs affecting whole or partial chromosomes, consistent with mosaicism reporting. Based on an expected LBR of 60% per transferred embryo in the euploid group and an absolute 5% difference in the putative mosaic group, along with a sampling rate of 0.16 (reflecting a 1:5 study-to-control allocation ratio) it was calculated that a total of 8,601 embryo transfers (7,415 euploid and 1,186 putative mosaic) would provide at least 90% power at a 5% significance level. This sample size was also adequately powered to assess miscarriage rates between the groups, assuming clinical miscarriage rates of 11% for euploid embryos and 14% for putative mosaics. Sample sizes were calculated by applying the formula for two Binomial Proportions reported in (Rosner, 2010).

Mean and standard deviation were used to describe continuous data, while percentages and 95% CI were used for categorical variables. Odds ratios (OR) and adjusted ORs were also computed. Multivariate logistic models were performed to evaluate the association of ICN presence with primary and secondary endpoints, by adjusting for the following clinical features: female age at oocyte retrieval, female BMI recorded at the closest time of embryo transfer, day of biopsy, morphology, prior history of failed embryo transfer(s). Statistical association of ICN and other feature with the endpoints was assessed within the logistic models by means of the Wald z-test. For each model, ORs adjusted for all the factors included were reported with a 95% CI. Significance of association was accepted with p < 0.05.

To assess the predictive value of clinical features and putative mosaicism findings in embryo selection, a sensitivity analysis was performed using a Receiver Operating Characteristic (ROC) curve, whereby the dataset was randomly split into training (70%) and test (30%) sets. Significance of the difference between AUCs of two ROC curves was computed with the Delong test.

To further understand the contribution of each factor and how different mosaic categories and/or levels explain the primary outcome, a decision tree model was fit on the training set. Within the pruning step, a complexity parameter (CP) was estimated in a cross-fold validation setting. The CP value corresponding to the minimum prediction error was chosen to control the size of the tree and to avoid overfitting.

Statistical analyses were performed using Python (version 3.10.8) with the packages pandas, numpy, scipy.stats, statsmodels, and scikit-learn. Data preprocessing and manipulation were conducted using pandas and numpy. Hypothesis testing and inferential statistics were performed with scipy.stats and statsmodels, while multivariate logistic models and sensitivity analyses implemented using scikit-learn. R (version 4.3.1) with packages rpart and rpart.plot was used to infer and visualize decision trees. The R library pROC was used to compute Delong test between pairs of ROC curves.

## Results

### Description of the study cohorts

This study includes a primary U.S. cohort of 6,951 patients who underwent 7,564 IVF cycles followed by 9,828 single transfers of embryos negative for WCA and SA following blastocyst-stage PGT-A. After unblinding the ICN status post-transfer, 84.7% (n = 8,328) of embryos were negative for ICN (euploid), while 8.8% (n = 864) exhibited sICN, and 5.6% (n = 562) displayed wcICN (Figure 1A). An additional small proportion (0.9%, n = 84) showed both sICN and wcICN. While embryos with both sICN and wcICN were excluded from the study analysis, clinical transfer outcomes were recorded (Supplementary Table 2).

**Figure 1:**
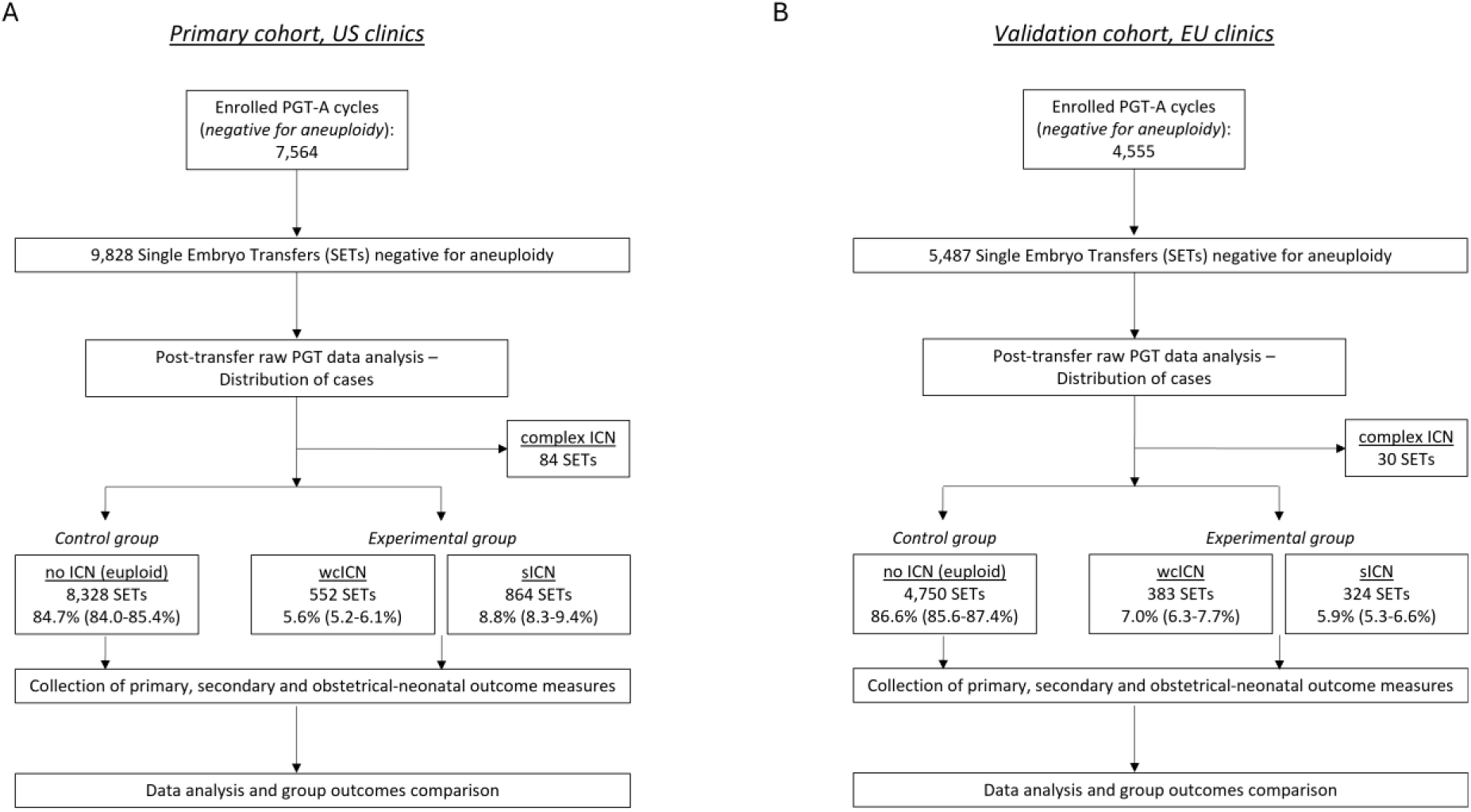
Study flowchart. (A) primary U.S. cohort analysis, (B) European (EU) validation cohort analysis. PGT-A=preimplantation genetic testing for aneuploidy; wcICN=whole chromosome intermediate copy number (ICN); sICN=segmental ICN; complex ICN=combination of wcICN and sICN.

The validation European cohort of 4,382 patients who underwent 4,555 IVF cycles and 5,487 single euploid embryo transfers followed the same study conditions but in a different IVF setting as shown in Figure 1B. Baseline demographic and cycle characteristics of the two study cohorts are summarized in Table 1. As anticipated due to differing PGT selection criteria between the two clinical practices, significant differences were observed between the cohorts in baseline patient demographics and clinical characteristics. In general, women in the U.S. cohort were younger (35.0 vs. 36.5 years, p<0.001) and had a more favorable prognosis. In fact, as expected, the U.S. data showed a higher number of retrieved oocytes and embryos per cycle, culminating in a higher number of euploid embryos and a higher live birth rate per transferred embryo, consistent with known trends. Slight procedural variations were identified, resulting in a higher proportion (57.9% vs. 25.5%, p<0.001) of embryos biopsied on day 6 in the U.S. cohort compared to the European cohort. Despite these differences, the overall putative mosaicism rate was found to be comparable between the two genetic testing sites (Table 1). Both cohorts corroborated the known positive association between advancing maternal age and aneuploidy rates and the lack of association of maternal age with ICN rate, further validating the reliability of datasets from the three genetic testing sites (Supplementary Figure 2 A/B).

**Table 1:**
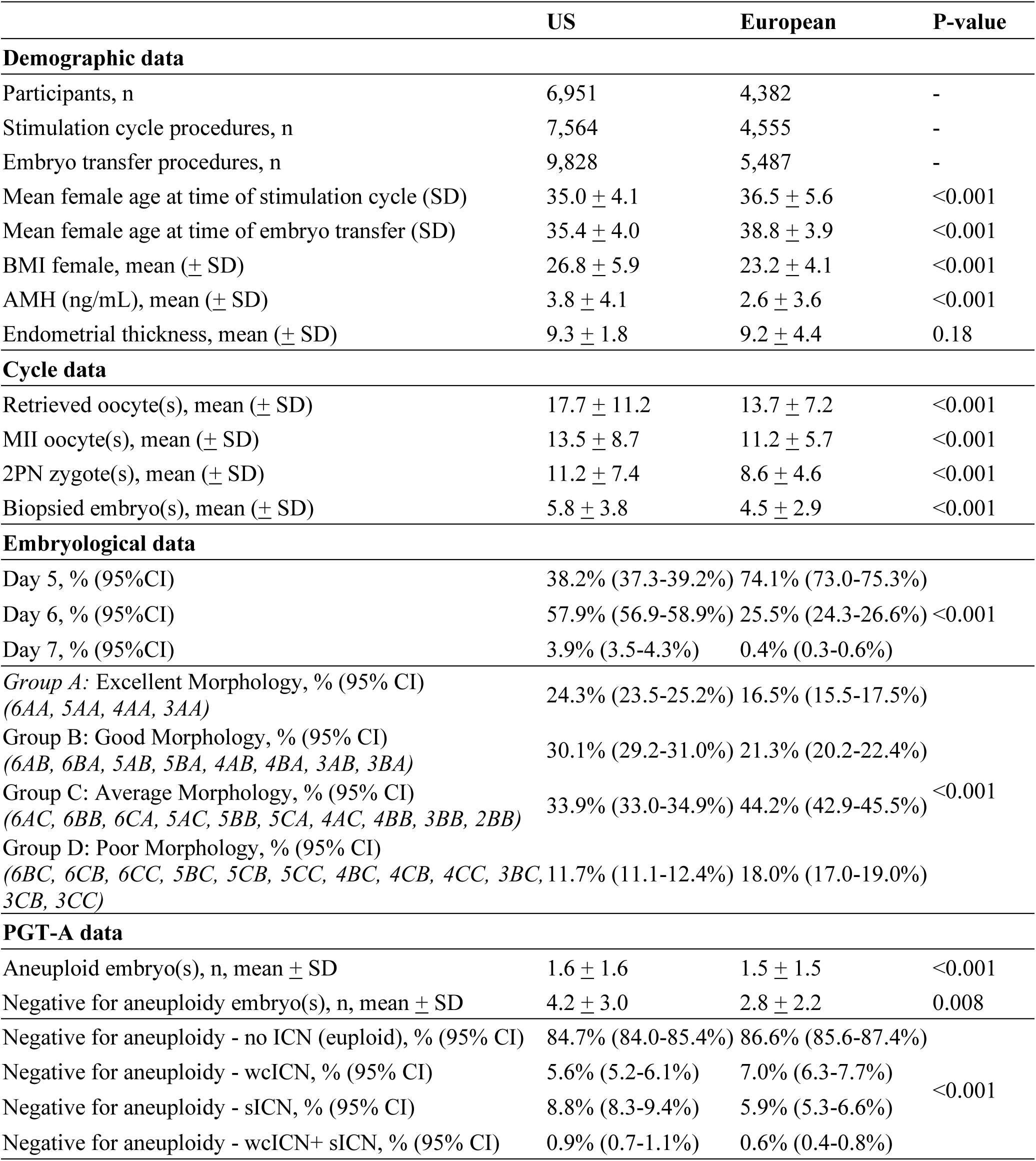
Baseline demographic, embryological and genetic data of cycles included in the study. BMI=body mass index; AMH=anti-müllerian hormone; MII=metaphase II; 2PN=two-pronuclear; PGT-A=preimplantation genetic testing for aneuploidy; wcICN=whole chromosome intermediate copy number (ICN); sICN=segmental ICN; complex ICN=combination of wcICN and sICN; SD=standard deviation.

### The primary U.S. cohort analysis

The analytical consistency of the PGT-A assay in detecting 10% incremental variations in intermediate copy number (ICN) was validated through cell line mixing experiments and demonstrated high reliability for both mosaicism losses and gains (Supplementary Table 1).

In the primary clinical cohort of putative mosaic embryo transfers, ICN was identified across all autosomes, without chromosome specific signatures as was seen for full aneuploidies in sibling embryos (Supplementary Figure 3A). To ensure that technical noise did not influence ICN calling, a metric evaluating the signal to noise ratio of copy number estimates (MAPD; Median Absolute Pairwise Difference) was assessed. No correlation was observed between MAPD and the maximum ICN deviation (Pearson R coefficient=0.117; Supplementary Figure 3B) suggesting independence from technical noise in the reported ICNs in the dataset.

After defining that ICN calling was independent of chromosome number and signal to noise ratio, its influence on clinical pregnancy outcomes was assessed. Logistic regression analysis demonstrated that the presence of ICN was associated with a slightly but statistically significant lower LBR (P=0.0001, OR=0.79, 95% CI: 0.7-0.89), as a consequence of the lower implantation and clinical pregnancy rates (Table 2). However, ICN was not associated with miscarriage rate (P=0.4893, OR=1.08, 95% CI=0.87-1.34). The evaluated covariates - including embryo morphology, day of biopsy, oocyte provider age, BMI, and history of previous embryo transfer failures—were also found to be independently associated with LBR (Figure 2A).

**Table 2:** Pregnancy, obstetrical and neonatal outcomes following the transfer of embryos negative for aneuploidy in the primary U.S. cohort.

| Pregnancy outcomes per transfer | no ICN (euploid) | ICN (wcICN or sICN) | Adjusted OR (95%CI, p value) | wcICN | Adjusted OR (95%CI, p value) | sICN | Adjusted OR (95%CI, p value) |
| --- | --- | --- | --- | --- | --- | --- | --- |
| Positive pregnancy test, % (n) | 78.7% (6553/8328) | 74.2% (1050/1416) | 0.84 (0.73-0.96, **0.0117**) | 70.7% (390/552) | 0.77 (0.63-0.94, **0.0114**) | 76.4% (660/864) | 0.89 (0.75-1.05, 0.173) |
| Biochemical pregnancy loss, % (n) | 10.7% (893/8328) | 13.0% (184/1416) | 1.22 (1.03-1.45, **0.0247**) | 15.2% (84/552) | 1.4 (1.09-1.8, **0.0082**) | 11.6% (100/864) | 1.1 (0.88-1.38, 0.376) |
| Clinical pregnancy, % (n) | 67.9% (5660/8328) | 61.2% (866/1416) | 0.79 (0.7-0.89, **0.0002**) | 55.4% (306/552) | 0.69 (0.57-0.82, **0.0001**) | 64.8% (560/864) | 0.87 (0.75-1.01, 0.073) |
| Miscarriage, % (n) | 7.6% (635/8328) | 7.9% (112/1416) | 1.08 (0.87-1.34, 0.4893) | 7.1% (39/552) | 0.98 (0.7-1.38, 0.9055) | 8.4% (73/864) | 1.14 (0.88-1.48, 0.316) |
| Live birth, % (n) | 60.0% (4993/8328) | 53.2% (753/1416) | 0.79 (0.7-0.89, **0.0001**) | 48.4% (267/552) | 0.71 (0.59-0.85, **0.0002**) | 56.2% (486/864) | 0.85 (0.73-0.98, 0.025) |
| **Obstetrical and neonatal outcomes** |  |  |  |  |  |  |  |
| Gestational hypertensive disorders, % (n) | 13.0% (655/5048) | 15.2% (115/756) | 1.17 (0.93-1.47, 0.1693) | 13.4% (36/268) | 0.97 (0.67-1.41, 0.86) | 16.2% (79/488) | 1.30 (0.997-1.683, 0.053) |
| Gestational diabetes, % (n) | 7.8% (393/5048) | 7.7% (58/756) | 0.99 (0.74-1.33, 0.9421) | 9.7% (26/268) | 1.23 (0.80-1.88, 0.34) | 6.6% (32/488) | 0.83 (0.57-1.22, 0.34) |
| Placental abruption, % (n) | 0.7% (36/5048) | 0.8% (6/756) | 0.96 (0.37-2.47, 0.9306) | 0.4% (1/268) | 0.49 (0.07-3.59, 0.48) | 1.0% (5/488) | 1.49 (0.58-3.84, 0.41) |
| Preterm delivery, % (n) | 10.7% (538/5048) | 10.4% (79/756) | 0.98 (0.76-1.27, 0.8947) | 9.0% (24/268) | 0.82 (0.53-1.27, 0.37) | 11.2% (55/488) | 1.07 (0.79-1.44, 0.68) |
| Low birth weight, % (n) | 6.2% (311/5048) | 8.1% (61/756) | 1.31 (0.97-1.77, 0.0815) | 7.1% (19/268) | 1.18 (0.73-1.92, 0.49) | 8.6% (42/488) | 1.41 (1.001-1.996, 0.050) |
| NICU admission, % (n) | 7.0% (351/5048) | 5.8% (44/756) | 0.82 (0.58-1.14, 0.2398) | 7.5% (20/268) | 1.10 (0.68-1.76, 0.70) | 4.9% (24/488) | 0.68 (0.44-1.05, 0.08) |
| Congenital anomalies, % (n) | 1.3% (65/5048) | 1.5% (11/756) | 1.19 (0.62-2.27, 0.6069) | 2.2% (6/268) | 1.55 (0.66-3.66, 0.31) | 1.0% (5/488) | 0.79 (0.32-1.97, 0.61) |

**Figure 2.**
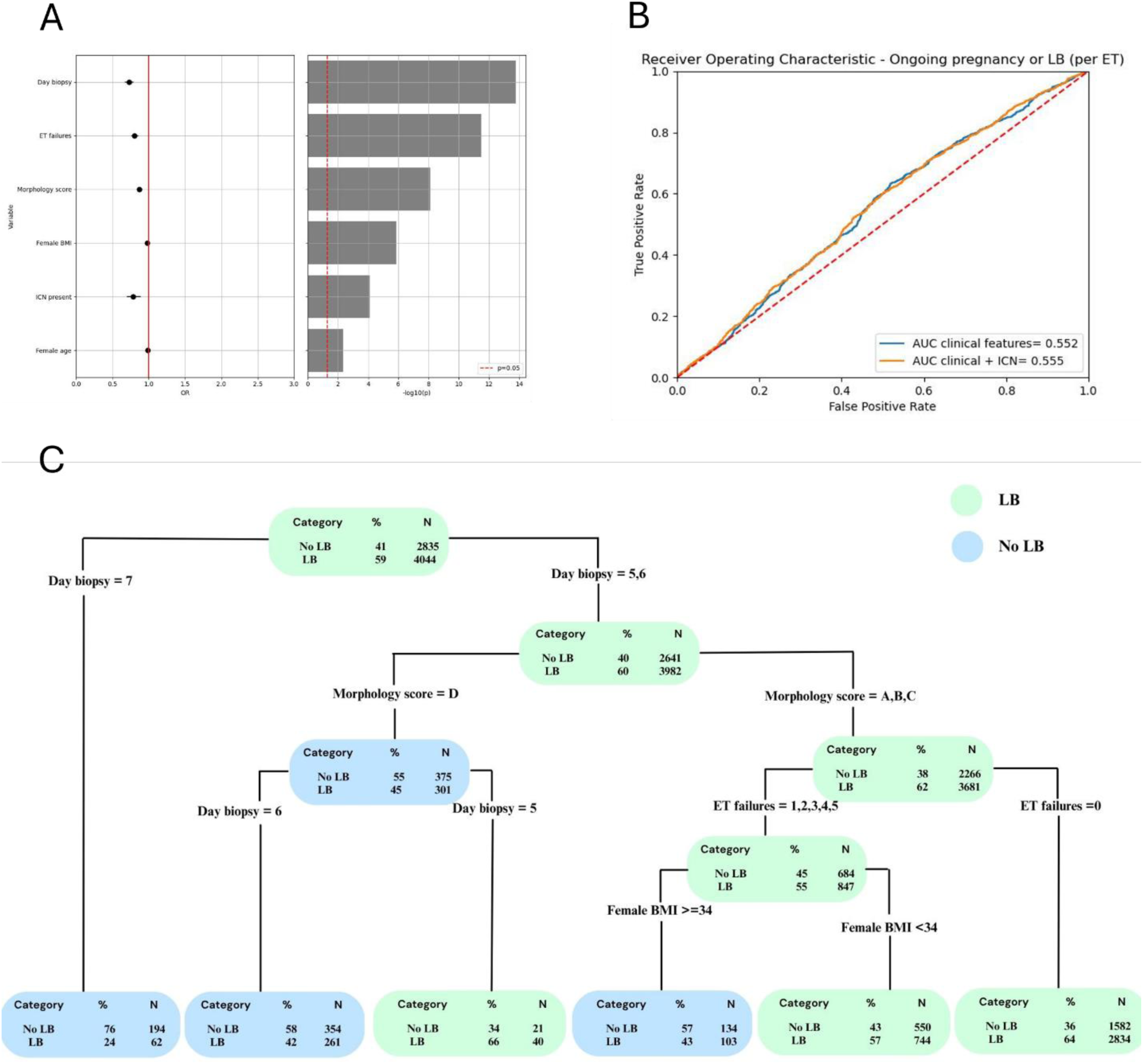
Association of ICN with LBR: primary U.S. cohort analysis. (A) Logistic regression analysis on the live birth rate for all relevant covariates. The left side displays the odds ratio (OR), representing the statistical effect, while the right side indicates the nominal statistical significance of each covariate. (B) ROC curve analysis of live birth rate prediction when including or excluding mosaicism status alongside established clinical features in PGT-A cycles. The analysis shows that although putative mosaicism alone was statistically associated with a slightly lower live birth rate (LBR), it was not found to enhance embryo selection or clinical decision-making when considered alongside stronger clinical predictors. This is likely due to the low overall incidence of putative mosaicism in embryos and its small effect size. (C) Decision tree model incorporating all clinical predictors from panel A. This analysis uses all covariates to identify the optimal embryo selection strategy across the full range of clinical scenarios represented in the dataset. Among embryos negative for uniform aneuploidies, the most influential parameters identified for prioritization were the day of biopsy and morphological grading. Notably, putative mosaicism (i.e., findings of wcICN or sICN) was not found to contribute additional predictive value and was not utilized by the model to guide embryo selection decisions.

It was then investigated whether a specific threshold in the ICN deviation values could be established as predictive of LB outcome. ICN deviation ranging from 15% to 89% was assessed as a continuous variable to identify whether an optimal separation could be identified between low and high ICN. By assuming a discrete increase of 10%, ICN deviation equal to or greater than 50% achieved nominal statistical significance (Supplementary Figure 4). The dataset was therefore divided into low (<50%) and high (>50%) ICN; based on this, 3.8% of transferred embryos had high ICN (N=377, high-wcICN=150 and high-sICN=277) while the remainder had low ICN. Occurrence of low and high ICN categories and relative outcomes are reported in Supplementary Table 3 and 4. Logistic regression analysis revealed that only high ICN values were statistically associated with a slightly lower LBR (P=0.02, OR=0.64, 95% CI=0.44-0.94). This effect was noted for both high wcICN and high sICN (Supplementary Figure 4).

Multivariate logistic models were performed to evaluate the association of ICN presence with secondary endpoints, by adjusting for the following clinical features: female age at oocyte retrieval, female BMI at embryo transfer, day of biopsy, morphology, prior history of failed embryo transfer(s). This analysis revealed no significant association between either wcICN or sICN and obstetrical and neonatal outcomes, neither maternal pathologies nor congenital anomalies of the fetus (Table 2). Individual chromosome level analysis for autosomes and sex chromosomes was not performed because of the insufficient sample size and for technical reasons, respectively. However, descriptive statistics for individual chromosomes and live birth rate outcomes are reported in Supplementary Tables 5 (autosomes) and 6 (sex chromosomes).

Next, to assess the clinical utility of these findings for embryo selection, a predictive model incorporating clinical, embryological, and chromosomal mosaicism features was developed by training a logistic regression model on 70% of the dataset, randomly selected. AUROC analysis demonstrated that embryo selection predictive performance was not significantly influenced by ICN status. The ROC and the corresponding AUC (Area Under the Curve) obtained when selecting embryos based solely on clinical and embryological factors (AUC = 0.552) was not significantly different from the one obtained when incorporating putative mosaicism findings in the model (AUC = 0.555; P > 0.05 for all comparisons) (Figure 2B). This indicates that, when evaluated alongside established clinical and embryological parameters, mosaicism had no clinical impact in predicting live birth among embryos that are negative for uniform chromosome aneuploidies. To assess whether any mosaic reporting strategy based on specific ICN type or level might support improved embryo selection, AUROC analyses were independently conducted for reporting of wc-ICN only; sICN only; high-wcICN only; high-sICN only; and both high wc-ICN and s-ICN cases (Supplementary Figure 5A-E). None of the ROCs and corresponding AUCs for any iteration of putative mosaic reporting were significantly different from the one based solely on clinical and embryological factors. The lack of predictive power of ICN is primarily attributable to (1) the low incidence of high ICN findings in the embryonic cohort (3.8%), combined with (2) the negligible effect size of ICN on live birth outcomes.

As an additional validation of these findings, a decision tree was fitted to better understand the contribution of each single clinical variable (ICN and others) in the prediction of LB (Figure 2C). Compared to the models previously reported, the decision tree enables the stratification of paths and values of each factor that lead to a positive or a negative outcome. In this model, factors that are not found to be adequately predictive of outcome are not retained after the pruning step of the model fitting. The retained factors are arranged in order of predictive value, from the root of the tree to the leaves. Remarkably, the model discarded ICN from the set of features that were informative for LB prediction, and thus ICN is not shown in the resulting tree. As expected, the tree automatically captured groups already known to be associated with a negative outcome (*e.g.,* day of biopsy = 7) or positive (*e.g.*, morphological groups = A, B, C).

### Clinical validation in independent clinical setting

To corroborate these findings in a different clinical setting—where the patient population undergoing IVF, as well as key clinical outcome measures, differ— a similar prospective blinded analysis was conducted for a validation group of blinded embryo transfers in European clinics. Specifically, an additional 5,487 single euploid embryo transfer cycles from multiple clinics in Spain were included (Figure 1B). Following unblinding of the PGT-A results, ICN rates for both wcICN and sICN were consistent with those observed in the U.S. dataset (Table 1).

The multivariate analysis of embryo transfer outcomes in the European setting confirmed all previously established associations (Supplementary Figure 6A, Supplementary Table 7, with the exception of body mass index (BMI). Notably, BMI distribution in the European cohort was narrower (Table 1) and less variable than in the U.S. population, potentially explaining this discrepancy. Similar predictive analyses conducted via ROC/AUC reaffirmed the lack clinical utility of mosaicism reporting when evaluated in conjunction with all clinical and laboratory features (Supplementary Figure 6B/C).

Importantly, in the European validation dataset, the absence of an association between ICN and adverse obstetrical and neonatal outcomes was again confirmed (Supplementary Table 7). However, it is important to note that not all pregnancy and neonatal outcomes recorded in the U.S. cohort were available for assessment in the European setting.

## Discussion

This study presents the most comprehensive, data-driven evaluation of the clinical impact of mosaicism reporting in PGT-A to date. By leveraging a uniquely large dataset and employing a blinded, non-selection approach, an unbiased assessment of the clinical significance of both whole chromosome and segmental ICN results - commonly reported as evidence of chromosomal mosaicism in embryos – was ensured. Furthermore, this study represents a novel effort to corroborate initial findings from a primary cohort with a validation cohort from a distinctly different IVF clinical setting. The consistency of ICN incidence and clinical outcomes across these two patient populations, despite differences in patient characteristics and clinical and embryological practices, demonstrates the robustness of the study findings.

Of great importance is the finding that the presence of ICN did not meaningfully impact clinical outcomes in a blinded setting, where putative mosaic embryos were not deliberately deprioritized. While embryos with high ICN deviations exhibited a slightly lower live birth rate, the effect was negligible when accounting for other embryological and clinical features. In fact, predictive models in both US and European cohorts, along with clinical decision trees evaluating optimal selection strategies, reinforced this finding: when multiple embryos negative for aneuploidy were available, against the existing backdrop of established and largely unmodifiable clinical features, mosaicism reporting does not result in higher live birth rate per transfer as compared to non-reporting of mosaicism. Additionally, clinical miscarriage rates, as well as obstetrical and neonatal outcomes following the transfer of embryos with ICN, were comparable to those of euploid embryos. This suggests that embryos with ICN detected by PGT-A at the blastocyst stage as a whole do not have increased obstetrical and fetal risks, negating the need for overly cautious patient counselling.

Collectively, these results therefore challenge the clinical relevance of widespread mosaicism reporting in PGT-A, questioning how and why this became clinical practice in the absence of prospective studies designed to assess the predictive value of any analytical metric used to diagnose mosaicism in preimplantation embryos. A recent survey conducted by the ESHRE Working Group on Chromosomal Mosaicism et al., 2022 reported that up to 15% of clinics recommended discarding or donating putative mosaic embryos for research as the primary course of action, while an additional 50% offered this approach as one of the available options for patients. Similar findings in multiple studies demonstrate that mosaicism reporting in PGT-A has significantly impacted embryo utilization policies (“Clinical Management of Mosaic Results from Preimplantation Genetic Testing for Aneuploidy of Blastocysts: A Committee Opinion,” 2023). It is not surprising therefore that reporting ICN findings in PGT-A as evidence of chromosomal mosaicism has significant implications for patients, notably unnecessarily increasing emotional distress and influencing clinical decision-making ultimately affecting embryo utilization policies.

While the reporting of mosaicism in PGT-A may have been initially warranted due to potential concerns over possible risks to pregnancy, the absence of supporting evidence after almost a decade of monitoring (Kahraman et al., 2020; Viotti et al., 2023), combined with the findings of this study, substantiates and reinforces the notion that ICN detected in PGT-A does not pose clinically significant reproductive nor neonatal risks and therefore should not be reported in the context of an evidence-based framework. Further compounding this are inconsistencies in ICN reporting criteria, which have contributed to variability in aneuploidy rates reported across PGT laboratories (Bardos et al., 2023; Popovic et al., 2024). This variability has diverted attention from the primary objective of PGT-A: identifying and de-selecting uniformly aneuploid embryos resulting from meiotic errors with well-established biological mechanisms. As a result, confusion among providers and patients has grown, leading to increasing skepticism about the overall benefit of aneuploidy testing in IVF-derived embryos in recent years.

Importantly, this study was not designed to elucidate the biological basis of ICN detected in TE biopsies but rather to rigorously assess its clinical relevance. While the findings here indicate that ICN reporting on this particular PGT-A platform has no clinical utility, a slight reduction in the live birth potential of embryos exhibiting high ICN levels for both whole and segmental aneuploidy was observed. Whether this reflects true mosaicism or is attributable to technical or analytical factors cannot be determined from this study, highlighting the need for further investigation into the nature of ICN in PGT-A. A previous study employing multifocal blastocyst biopsies using the same PGT-A platform have demonstrated low reproducibility of mosaic calls across different regions of the same embryo, especially when compared to the high confirmation rates observed for euploid and aneuploid diagnoses (J. Kim et al., 2022). To date, the largest rebiopsy studies have rarely confirmed the presence of pervasive mosaicism in blastocyst-stage embryos or provided mechanistic evidence to support its association to ICNs. Observations such as reciprocal chromosomal gains and losses involving the same chromosome—potentially indicative of mitotic errors— have been reported only in rare instances (Capalbo et al., 2021; J. Kim et al., 2022). Moreover, follow-up genetic analyses of products of conception and postnatal outcomes have consistently failed to corroborate mosaicism findings initially identified in TE biopsies (Huang et al., 2009; J. Kim et al., 2022; Treff & Marin, 2021). Although earlier follow-up at the stage of early developmental arrest post-transfer could provide further insights, this remains largely inaccessible for genetic investigation. However, experimental embryological models that extend in vitro embryo development to day 12 have not confirmed the presence of mosaicism originally detected in clinical TE biopsies (Popovic et al., 2019). Furthermore, recent evidence suggests that certain PGT platforms may systematically misclassify aneuploid embryos as mosaic (Popa et al., 2025). Collectively, these independent findings challenge the fundamental capability of PGT to accurately detect true and clinically meaningful mosaicism in embryos, further highlighting the importance of this study.

Certain limitations of this study must be acknowledged. Notably, the findings are specific to this PGT platform and may not be generalizable to other PGT-A methodologies. This underscores the need for prospective, blinded studies to assess the predictive value of findings, including putative mosaicism, across different PGT platforms prior to reporting them clinically. Data on prior embryo transfer failures were limited to records from referring clinics and did not include information on any previous reproductive treatments conducted elsewhere. Additionally, obstetrical and neonatal outcomes in this study were self-reported, introducing the potential for reporting bias. To mitigate this, follow-up pregnancy data were all collected by experienced clinicians to ensure consistency and accuracy. Furthermore, while this analysis is based on an unprecedented sample size, the study lacks sufficient power to explore chromosome-specific associations between ICNs and clinical outcomes. Larger studies than the one presented here are needed to determine whether specific combinations of chromosomes and ICN types have a distinct impact on clinical outcomes.

Despite these limitations, this study has several notable strengths. It employs a blinded, non-selection study design, ensuring an unbiased evaluation of clinical outcomes, and the use of a validated PGT-A platform for detecting whole chromosome aneuploidies (utilizing the same blinded, non-selection study design) enhances the reliability of the findings. Moreover, the study benefits from a large sample size, providing robust statistical power, and includes various types of ICN, allowing for a comprehensive assessment. The inclusion of an independent validation cohort from a clinically diverse patient population and IVF practice, further strengthens the clinical generalizability of the findings. The observed differences in clinical context and IVF protocols between the US and EU cohorts were both expected and necessary, providing a valuable opportunity to assess the applicability of PGT-A mosaicism reporting across varied clinical environments. This, in turn, reinforces the robustness of the conclusions drawn here. Furthermore, the consistency in identifying similar rates of aneuploidy and intermediate ICN across the two datasets underscores the high degree of standardization and uniformity in genetic laboratory practices between the two testing sites. Despite serving distinct clinical IVF practices, this demonstrated reproducibility in the PGT-A findings, reinforcing the reliability of the methodologies employed in this study.

For nearly a decade, the observation of an ICN in PGT has been interpreted as evidence of mosaicism and poorer reproductive outcomes without proper validation, fueling substantial controversy and uncertainty in both clinical and research settings, and ultimately influencing embryo clinical utilization policies. This study provides strong evidence that ICN findings detected by PGTSeq do not enable enhanced embryo assessment or selection decisions that improve patient outcomes, even with the support of SNP-based genotyping data. The reporting of putative mosaicism lacks clinical utility and should not influence embryo selection policies moving forward. When considered alongside prior blinded research evaluating the clinical predictive value of uniform whole chromosome aneuploidies on embryonic reproductive competence (Tiegs et al., 2021), the findings collectively support that the only evidence-based application of PGT-A is the deselection of embryos with uniform aneuploidies. As is widely recognized in clinical genetics, testing should be evidence-based – the only findings reported to clinicians and patients being those having demonstrable clinical validity, confirmed in rigorous studies (Deans et al., 2022; Richards et al., 2015; Smith et al., 2015). The unnecessary harm caused by widespread mosaicism reporting in PGT-A prior to assessment of its value in appropriately designed studies should serve as a critical lesson for the field, shaping future developments as new genomic technologies emerge. As PGT technology advances, particularly in the context of genome sequencing, it is imperative that robust frameworks are established to ensure that only accurate and clinically relevant genetic findings are reported. By adopting a more evidence-based approach, the field can prioritize patient safety and outcomes while avoiding the pitfalls of premature clinical implementation.

## Author role

AC and CJ conceived and oversaw the study. PG, XT, YZ, FM, CSO, LP, and SC conducted data collection and analysis. DB, GC, EFM, CM, and VJ performed laboratory testing. MW, RS, TM, JP, VVB, ARM, AP, and JGV supervised clinical practice, patient recruitment, and counseling throughout the study period. EM critically reviewed the data and contributed to manuscript drafting. All authors provided constructive feedback on data analysis and manuscript development.

## Supporting information

manuscript_

## Data Availability

All data produced in the present study are available upon reasonable request to the authors

## References

1. Bardos, J., Kwal, J., Caswell, W., Jahandideh, S., Stratton, M., Tucker, M., DeCherney, A., Devine, K., Hill, M., & O’Brien, J. E. (2023). Reproductive genetics laboratory may impact euploid blastocyst and live birth rates: a comparison of 4 national laboratories’ PGT-A results from vitrified donor oocytes. Fertility and Sterility, 119(1). 10.1016/j.fertnstert.2022.10.010

2. Capalbo, A., Poli, M., Jalas, C., Forman, E. J., & Treff, N. R. (2022). On the reproductive capabilities of aneuploid human preimplantation embryos. In American Journal of Human Genetics (Vol. 109, Issue 9). 10.1016/j.ajhg.2022.07.009

3. Capalbo, A., Poli, M., Rienzi, L., Girardi, L., Patassini, C., Fabiani, M., Cimadomo, D., Benini, F., Farcomeni, A., Cuzzi, J., Rubio, C., Albani, E., Sacchi, L., Vaiarelli, A., Figliuzzi, M., Findikli, N., Coban, O., Boynukalin, F. K., Vogel, I., … Simón, C. (2021). Mosaic human preimplantation embryos and their developmental potential in a prospective, non-selection clinical trial. American Journal of Human Genetics, 108(12). 10.1016/j.ajhg.2021.11.002

4. Capalbo, A., Ubaldi, F. M., Rienzi, L., Scott, R., & Treff, N. (2017). Detecting mosaicism in trophectodermbiopsies: Current challenges and future possibilities. In Human Reproduction (Vol. 32, Issue 3). 10.1093/humrep/dew250

5. Clinical management of mosaic results from preimplantation genetic testing for aneuploidy of blastocysts: a committee opinion. (2023). Fertility and Sterility, 120(5). 10.1016/j.fertnstert.2023.08.969

6. Deans, Z. C., Ahn, J. W., Carreira, I. M., Dequeker, E., Henderson, M., Lovrecic, L., Õunap, K., Tabiner, M., Treacy, R., & van Asperen, C. J. (2022). Recommendations for reporting results of diagnostic genomic testing. European Journal of Human Genetics, 30(9). 10.1038/s41431-022-01091-0

7. ESHRE Working Group on Chromosomal Mosaicism, De Rycke, M., Capalbo, A., Coonen, E., Coticchio, G., Fiorentino, F., Goossens, V., Mcheik, S., Rubio, C., Sermon, K., Sfontouris, I., Spits, C., Vermeesch, J. R., Vermeulen, N., Wells, D., Zambelli, F., & Kakourou, G. (2022). ESHRE survey results and good practice recommendations on managing chromosomal mosaicism. Human Reproduction Open, 2022(4), hoac044. 10.1093/hropen/hoac044

8. Franasiak, J. M., Forman, E. J., Hong, K. H., Werner, M. D., Upham, K. M., Treff, N. R., & Scott, R. T. (2014). The nature of aneuploidy with increasing age of the female partner: A review of 15,169 consecutive trophectoderm biopsies evaluated with comprehensive chromosomal screening. Fertility and Sterility, 101(3). 10.1016/j.fertnstert.2013.11.004

9. Gardner, D., & Schoolcraft, W. (1999). In-vitro culture of human blastocysts. In Towards reproductive certainty: fertility and genetics.

10. Girardi, L., Figliuzzi, M., Poli, M., Serdarogullari, M., Patassini, C., Caroselli, S., Pergher, I., Cogo, F., Coban, O., Boynukalin, F. K., Bahceci, M., Navarro, R., Rubio, C., Findikli, N., Simón, C., & Capalbo, A. (2023). The use of copy number loads to designate mosaicism in blastocyst stage PGT-A cycles: fewer is better. *Human Reproduction (Oxford*, England), 38(5), 982–991. 10.1093/humrep/dead049

11. Goodrich, D., Tao, X., Bohrer, C., Lonczak, A., Xing, T., Zimmerman, R., Zhan, Y., Scott, R. T., & Treff, N. R. (2016). A randomized and blinded comparison of qPCR and NGS-based detection of aneuploidy in a cell line mixture model of blastocyst biopsy mosaicism. Journal of Assisted Reproduction and Genetics, 33(11). 10.1007/s10815-016-0784-3

12. Greco, E., Minasi, M. G., & Fiorentino, F. (2015). Healthy Babies after Intrauterine Transfer of Mosaic Aneuploid Blastocysts. New England Journal of Medicine, 373(21). 10.1056/nejmc1500421

13. Gruhn, J. R., Zielinska, A. P., Shukla, V., Blanshard, R., Capalbo, A., Cimadomo, D., Nikiforov, D., Chan, A. C.-H., Newnham, L. J., Vogel, I., Scarica, C., Krapchev, M., Taylor, D., Kristensen, S. G., Cheng, J., Ernst, E., Bjørn, A.-M. B., Colmorn, L. B., Blayney, M., … Hoffmann, E. R. (2019). Chromosome errors in human eggs shape natural fertility over reproductive life span. Science (New York, N.Y.), 365(6460), 1466–1469. 10.1126/science.aav7321

14. Hassold, T., Chen, N., Funkhouser, J., Jooss, T., Manuel, B., Matsuura, J., Matsuyama, A., Wilson, C., Yamane, J. A., & Jacobs, P. A. (1980). A cytogenetic study of 1000 spontaneous abortions. Annals of Human Genetics, 44(2). 10.1111/j.1469-1809.1980.tb00955.x

15. Hipp, H. S., Crawford, S., Boulet, S., Toner, J., Sparks, A. A. E., & Kawwass, J. F. (2022). Trends and Outcomes for Preimplantation Genetic Testing in the United States, 2014-2018. In JAMA (Vol. 327, Issue 13). 10.1001/jama.2022.1892

16. Huang, A., Adusumalli, J., Patel, S., Liem, J., Williams, J., & Pisarska, M. D. (2009). Prevalence of chromosomal mosaicism in pregnancies from couples with infertility. Fertility and Sterility, 91(6). 10.1016/j.fertnstert.2008.03.044

17. Kahraman, S., Cetinkaya, M., Yuksel, B., Yesil, M., & Cetinkaya, C. P. (2020). The birth of a baby with mosaicism resulting from a known mosaic embryo transfer: A case report. Human Reproduction, 35(3). 10.1093/humrep/dez309

18. Kim, J., Tao, X., Cheng, M., Steward, A., Guo, V., Zhan, Y., Scott, R. T., & Jalas, C. (2022). The concordance rates of an initial trophectoderm biopsy with the rest of the embryo using PGTseq, a targeted next-generation sequencing platform for preimplantation genetic testing-aneuploidy. Fertility and Sterility, 117(2). 10.1016/j.fertnstert.2021.10.011

19. Kim, T., Neblett, M. F., Shandley, L. M., Omurtag, K., Hipp, H. S., & Kawwass, J. F. (2018). Mosaic embryo transfer: a survey of current U.S. ART clinic practices. Fertility and Sterility, 110(4). 10.1016/j.fertnstert.2018.07.1168

20. Kung, A., Munné, S., Bankowski, B., Coates, A., & Wells, D. (2015). Validation of next-generation sequencing for comprehensive chromosome screening of embryos. Reproductive BioMedicine Online, 31(6). 10.1016/j.rbmo.2015.09.002

21. Mumusoglu, S., Telek, S. B., & Ata, B. (2025). Preimplantation genetic testing for aneuploidy in unexplained recurrent pregnancy loss: a systematic review and meta-analysis. Fertility and Sterility, 123(1), 121–136. 10.1016/j.fertnstert.2024.08.326

22. Munné, S., Blazek, J., Large, M., Martinez-Ortiz, P. A., Nisson, H., Liu, E., Tarozzi, N., Borini, A., Becker, A., Zhang, J., Maxwell, S., Grifo, J., Babariya, D., Wells, D., & Fragouli, E. (2017). Detailed investigation into the cytogenetic constitution and pregnancy outcome of replacing mosaic blastocysts detected with the use of high-resolution next-generation sequencing. Fertility and Sterility, 108(1). 10.1016/j.fertnstert.2017.05.002

23. Popa, T., Davis, C., Xanthopoulou, L., Bakosi, E., He, C., O’Neill, H., & Ottolini, C. S. (2025). Current quantitative methodologies for preimplantation genetic testing frequently misclassify meiotic aneuploidies as mosaic. Fertility and Sterility. 10.1016/j.fertnstert.2025.02.018

24. Popovic, M., Borot, L., Lorenzon, A. R., De Castro Lopes, A. L. R., Sakkas, D., Lledó, B., Morales, R., Ortiz, J. A., Polyzos, N. P., Parriego, M., Azpiroz, F., Galain, M., Pujol, A., Menten, B., Dhaenens, L., Meerschaut, F. Vanden, Stoop, D., Rodríguez, M., De La Blanca, E. P., … Vassena, R. (2024). Ímplicit bias in diagnosing mosaicismãmongst preimplantation genetic testing providers: results fromã multicenter study of 36 395 blastocysts. Human Reproduction, 39(1). 10.1093/humrep/dead213

25. Popovic, M., Dhaenens, L., Taelman, J., Dheedene, A., Bialecka, M., De Sutter, P., Chuva De Sousa Lopes, S. M., Menten, B., & Heindryckx, B. (2019). Extended in vitro culture of human embryos demonstrates the complex nature of diagnosing chromosomal mosaicism from a single trophectoderm biopsy. Human Reproduction, 34(4). 10.1093/humrep/dez012

26. Rana, B., Lambrese, K., Mendola, R., Xu, J., Garrisi, J., Miller, K., Marin, D., & Treff, N. R. (2023). Identifying parental and cell-division origins of aneuploidy in the human blastocyst. American Journal of Human Genetics, 110(4), 565–574. 10.1016/j.ajhg.2023.03.003

27. Richards, S., Aziz, N., Bale, S., Bick, D., Das, S., Gastier-Foster, J., Grody, W. W., Hegde, M., Lyon, E., Spector, E., Voelkerding, K., & Rehm, H. L. (2015). Standards and guidelines for the interpretation of sequence variants: a joint consensus recommendation of the American College of Medical Genetics and Genomics and the Association for Molecular Pathology. Genetics in Medicine, 17(5), 405–424. 10.1038/gim.2015.30

28. Rosner, B. (2010). Hypothesis Testing: Categorical Data. In Fundamentals of Biostatistics.

29. Sacchi, L., Albani, E., Cesana, A., Smeraldi, A., Parini, V., Fabiani, M., Poli, M., Capalbo, A., & Levi-Setti, P. E. (2019). Preimplantation Genetic Testing for Aneuploidy Improves Clinical, Gestational, and Neonatal Outcomes in Advanced Maternal Age Patients Without Compromising Cumulative Live-Birth Rate. Journal of Assisted Reproduction and Genetics, 36(12). 10.1007/s10815-019-01609-4

30. Smith, K., Martindale, J., Wallis, Y., Bown, N., Leo, N., Creswell, L., Fews, G., & Deans, Z. (2015). General Genetic Laboratory Reporting Recommendations. https://www.acgs.uk.com/media/10758/acgs_general_genetic_laboratory_reporting_recommendations_2015.pdf

31. Tiegs, A. W., Tao, X., Zhan, Y., Whitehead, C., Kim, J., Hanson, B., Osman, E., Kim, T. J., Patounakis, G., Gutmann, J., Castelbaum, A., Seli, E., Jalas, C., & Scott, R. T. (2021). A multicenter, prospective, blinded, nonselection study evaluating the predictive value of an aneuploid diagnosis using a targeted next-generation sequencing–based preimplantation genetic testing for aneuploidy assay and impact of biopsy. Fertility and Sterility, 115(3). 10.1016/j.fertnstert.2020.07.052

32. Treff, N. R., & Marin, D. (2021). The “mosaic” embryo: misconceptions and misinterpretations in preimplantation genetic testing for aneuploidy. In Fertility and Sterility (Vol. 116, Issue 5). 10.1016/j.fertnstert.2021.06.027

33. Ubaldi, F. M., Capalbo, A., Colamaria, S., Ferrero, S., Maggiulli, R., Vajta, G., Sapienza, F., Cimadomo, D., Giuliani, M., Gravotta, E., Vaiarelli, A., & Rienzi, L. (2015). Reduction of multiple pregnancies in the advanced maternal age population after implementation of an elective single embryo transfer policy coupled with enhanced embryo selection: Pre- and post-intervention study. Human Reproduction, 30(9). 10.1093/humrep/dev159

34. Verlinsky, Y., Ginsberg, N., Lifchez, A., Valle, J., Moise, J., & Strom, C. M. (1990). Analysis of the first polar body: preconception genetic diagnosis. *Human Reproduction (Oxford*, England), 5(7), 826–829. 10.1093/oxfordjournals.humrep.a137192

35. Viotti, M., Greco, E., Grifo, J. A., Madjunkov, M., Librach, C., Cetinkaya, M., Kahraman, S., Yakovlev, P., Kornilov, N., Corti, L., Biricik, A., Cheng, E. H., Su, C. Y., Lee, M. S., Bonifacio, M. D., Cooper, A. R., Griffin, D. K., Tran, D. Y., Kaur, P., … Spinella, F. (2023). Chromosomal, gestational, and neonatal outcomes of embryos classified as a mosaic by preimplantation genetic testing for aneuploidy. Fertility and Sterility, 120(5). 10.1016/j.fertnstert.2023.07.022

36. Zegers-Hochschild, F., Adamson, G. D., Dyer, S., Racowsky, C., de Mouzon, J., Sokol, R., Rienzi, L., Sunde, A., Schmidt, L., Cooke, I. D., Simpson, J. L., & van der Poel, S. (2017). The International Glossary on Infertility and Fertility Care, 2017. Fertility and Sterility, 108(3), 393–406. 10.1016/j.fertnstert.2017.06.005

