## Supplementary material for "PGT-A Mosaicism Reporting Lacks Clinical Predictive Value: Evidence from a Multisite, Double-Blinded Study with Independent Validation Across 15,315 Single-Embryo Transfers": manuscript_

**Supplementary Figure 1: Aneuploidy Detection and Reporting in the PGTSeq Assay**

In the PGTSeq assay, the determination and reporting of aneuploidies relies on the combined analysis of chromosomal copy number and the alternate allele frequency (B allelic frequency, or BAF) computed from SNPs.
For monosomies, a reduced copy number consistent with a single chromosome copy, along with loss of heterozygosity (LOH) across the chromosome (BAF consistent with only A or B configurations), supports the diagnosis of a uniform chromosomal loss (**Figure A and B for uniform whole chromosome and segmental losses**, respectively). In contrast, when a reduced intermediate copy number (ICN) is observed but heterozygous SNPs are still detected—as indicated by a positive B-allelic frequency (BAF)—the result is interpreted as a mosaic loss (**Figure C and D for mosaic whole chromosome and segmental losses**, respectively). In such cases, the sample is reported as negative for uniform aneuploidy.
For trisomies, a copy number consistent with three chromosomal copies, along with a split in the BAF (as expected in AAB or ABB configurations), confirms a chromosomal gain (**Figure E and F for uniform whole chromosome and segmental gains**, respectively). In contrast, when a gained ICN is detected without a corresponding full deviation in the BAF from the expected ratio for disomic heteroparental pair of chromosomes, a uniform abnormality cannot be confirmed, and the result is interpreted as a mosaic gain. In such cases, the sample is reported as negative for uniform aneuploidy (**Figure G and H for mosaic whole chromosome and segmental gains**, respectively).
These interpretation and reporting criteria were developed and validated through a prospective blinded trial (Tiegs et al.2021), cell line mixing experiments (detailed below), and extensive internal audits and QC assessments, demonstrating over 99% concordance between manual clinical laboratory reviews and the automated calls generated by the PGTSeq software.

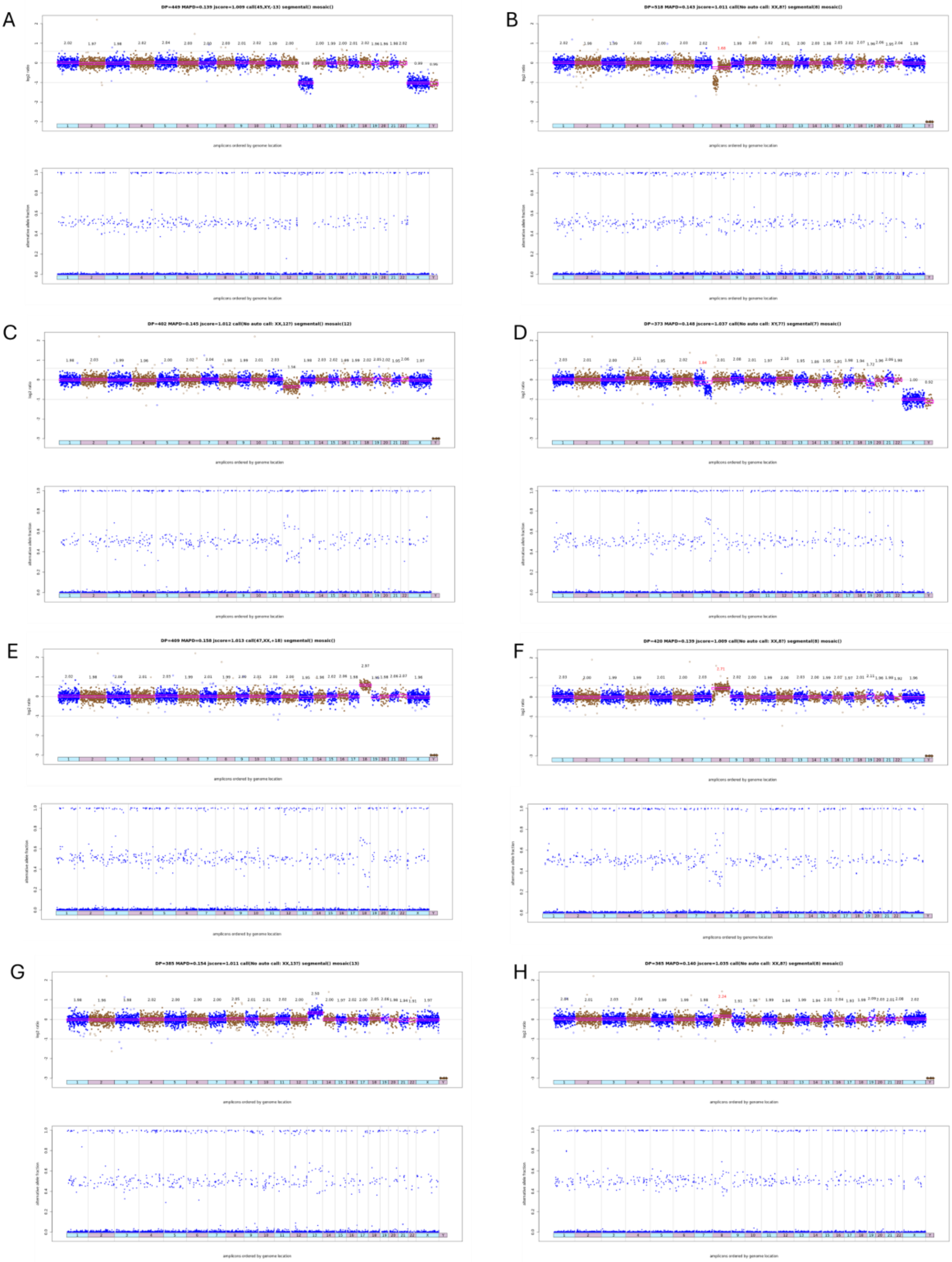
A
B
C
D
E
F
G
H

**Supplementary Figure 2:** (A) aneuploidy and ICN patterns per female age in primary U.S. cohort, (B) aneuploidy and ICN patterns per female age in validation European cohort, (C) aneuploidy patterns per chromosome in primary U.S. cohort.

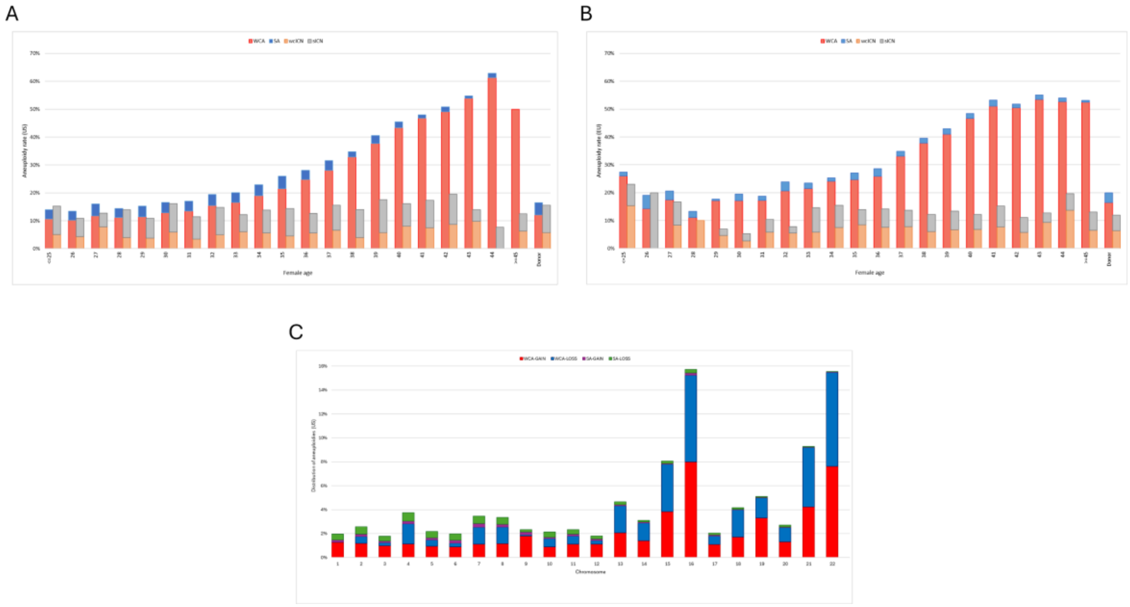


**Supplementary Figure 3.** Distribution of (A) wcICNs and (B) sICNs per chromosome in primary U.S. cohort after unblinding and their correlation with MAPD (C).

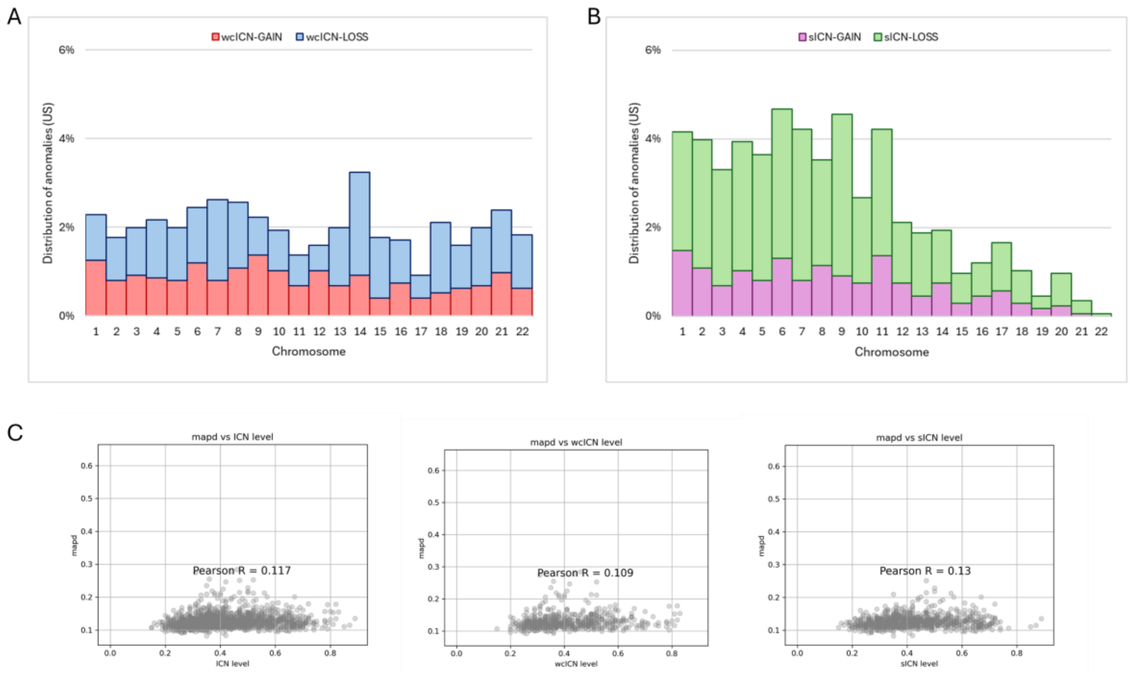

**Supplementary Figure 4: Association of ICN with LBR in primary U.S. cohort analysis** according to low and high grade with a threshold of 50% as maximum ICN deviation from the standard disomy state (A). Outcomes are further stratified by 10% increments in intermediate copy number (ICN) for both segmental (B) and whole-chromosome (C) variations.

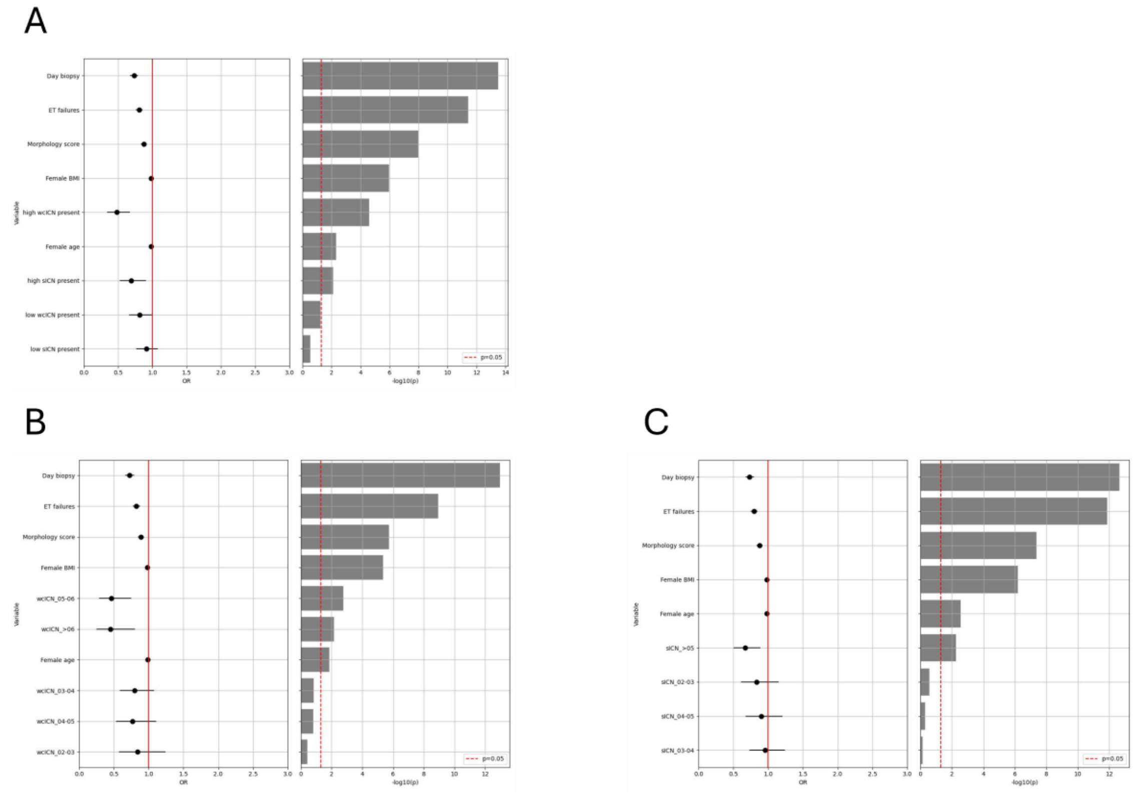

**Supplementary Figure 5: Impact of ICN reporting on LBR in primary U.S. cohort; analysis based on ROC analysis,** for (A) wcICN; (B) sICN; (C) high- wcICN only, (D) high-sICN only, (E) high wcICN and high-sICN combined.

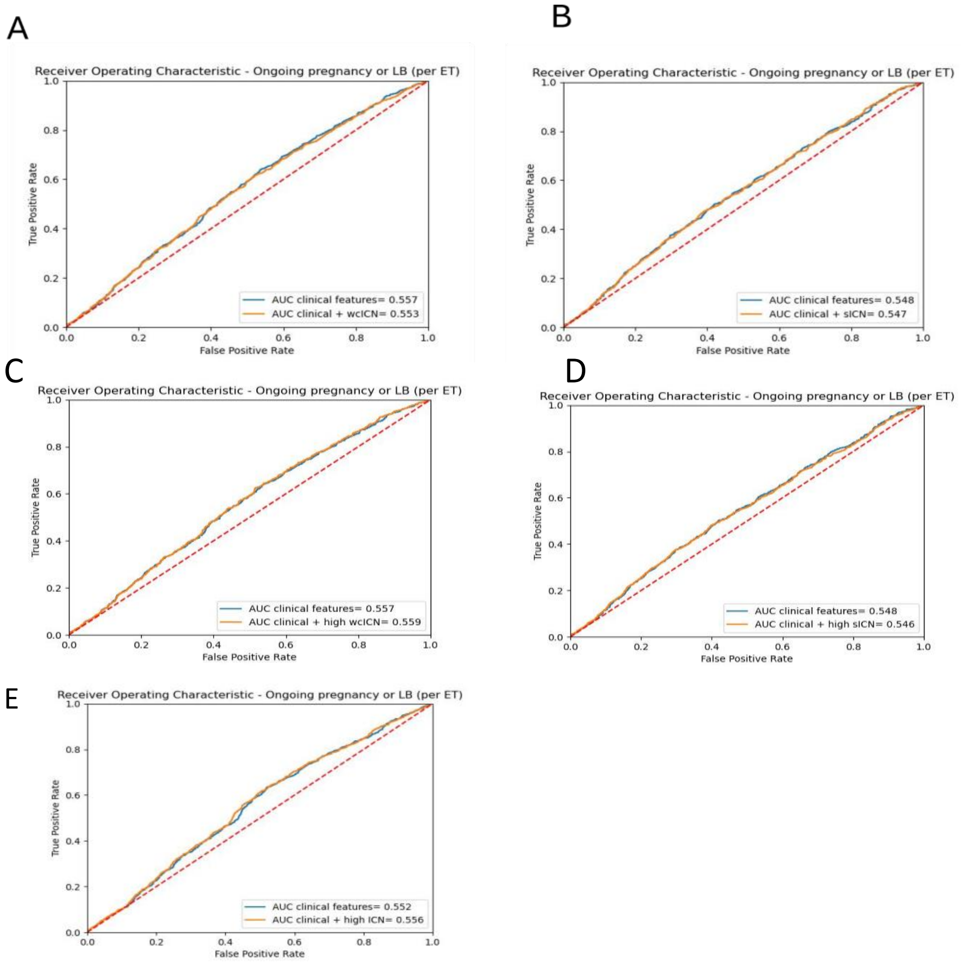

**Supplementary Figure 6. Association of ICN with LBR in the validation European cohort analysis.** (A) Logistic regression analysis including covariates. (B) ROC curve. (C) Decision tree.

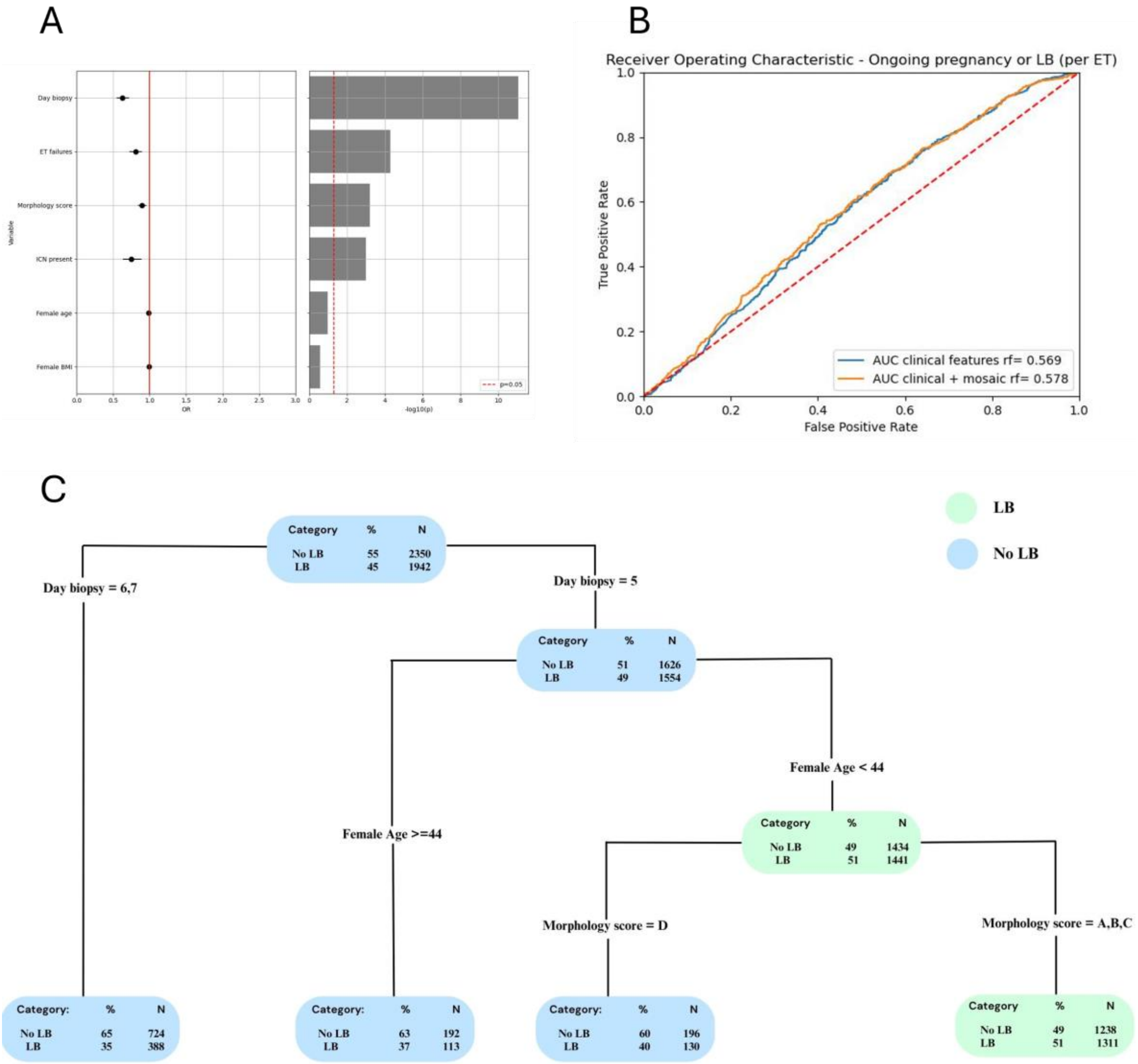

**Supplementary Table 1. ICN validation**. Experimental mixtures of cell lines (GM01359 carrying 47,XY,+18 and GM02948 carrying 47,XY,+13) at different proportions to mimic chromosomal mosaicism of different levels. CN=copy number value, SD=standard deviation, wcICN-GAIN=intermediate copy number of a whole chromosome towards trisomy.

| ID Sample | Proportion for the cell line mixtures used (chr18 : chr13) | chr18 (CN ± SD) | chr13 (CN ± SD) | Interpretation |
|---|---|---|---|---|
| 1 | 10 : 0 | 3.01 ± 0.02 | 2.01 ± 0.00 | Trisomy chr18 |
| 2 | 9 : 1 | 2.88 ± 0.04 | 2.10 ± 0.01 | wcICN – GAIN (chr18) |
| 3 | 8 : 2 | 2.78 ± 0.03 | 2.22 ± 0.06 | wcICN – GAIN (chr18) |
| 4 | 7 : 3 | 2.70 ± 0.02 | 2.32 ± 0.05 | wcICN – GAIN (chr18) |
| 5 | 6 : 4 | 2.59 ± 0.04 | 2.41 ± 0.03 | wcICN – GAIN (chr18) |
| 6 | 5 : 5 | 2.51 ± 0.06 | 2.52 ± 0.04 | - |
| 7 | 4 : 6 | 2.39 ± 0.05 | 2.59 ± 0.04 | wcICN – GAIN (chr13) |
| 8 | 3 : 7 | 2.26 ± 0.02 | 2.74 ± 0.06 | wcICN – GAIN (chr13) |
| 9 | 2 : 8 | 2.20 ± 0.01 | 2.78 ± 0.09 | wcICN – GAIN (chr13) |
| 10 | 1 : 9 | 2.11 ± 0.03 | 2.90 ± 0.06 | wcICN – GAIN (chr13) |
| 11 | 0 : 10 | 2.00 ± 0.04 | 3.03 ± 0.06 | Trisomy chr13 |

**Supplementary Table 2.** Pregnancy outcomes for embryos exhibiting a combination of wcICN and sICN (i.e., complex ICN) in primary U.S. cohort.

| | complex ICN |
|---|---|
| Positive pregnancy test, % (n) | 69.0% (58/84) |
| Biochemical pregnancy loss, % (n) | 14.3% (12/84) |
| Clinical pregnancy, % (n) | 54.8% (46/84) |
| Miscarriage, % (n) | 8.3% (7/84) |
| Ongoing/Live birth, % (n) | 46.4% (39/84) |

**Supplementary Table 3:** Pregnancy outcomes for embryos exhibiting low and high ICN based on a threshold of 50% as maximum ICN deviation from the standard two copies. Distributions are calculated in primary U.S. cohort, not reporting data for complex ICN (0.9%, n=84/9828).

| | **Positive pregnancy test**, % (n) | **Biochemical pregnancy loss**, % (n) | **Clinical pregnancy**, % (n) | **Miscarriage**, % (n) | **Ongoing/Live birth**, % (n) |
|---|---|---|---|---|---|
| **Euploid (no ICN**; 84.7%; n=8328/9828) | 78.7% (6553/8328) | 10.7% (893/8328) | 68.0% (5660/8328) | 7.8% (649/8328) | 60.2% (5011/8328) |
| ICN (10.6%; n=1040/9828) | 75.6% (786/1040) | 11.9% (124/1040) | 63.7% (662/1040) | 8.1% (84/1040) | 55.6% (578/1040) |
| wcICN (4.1%; n=403/9828) | 72.7% (293/403) | 15.4% (62/403) | 57.3% (231/403) | 6.2% (25/403) | 51.1% (206/403) |
| sICN (6.5%; n=637/9828) | 77.4% (493/637) | 9.7% (62/637) | 67.7% (431/637) | 9.3% (59/637) | 58.4% (372/637) |
| ICN (3.8%; n=376/9828) | 70.2% (264/376) | 16.0% (60/376) | 54.3% (204/376) | 7.7% (29/376) | 46.5% (175/376) |
| wcICN (1.5%; n=149/9828) | 65.1% (97/149) | 14.8% (22/149) | 50.3% (75/149) | 9.4% (14/149) | 40.9% (61/149) |
| sICN (2.3%; n=227/9828) | 73.6% (167/227) | 16.7% (38/227) | 56.8% (129/227) | 6.6% (15/227) | 50.2% (114/227) |

**Supplementary Table 4:** Live birth outcomes of embryos negative for aneuploidy and positive for whole chromosome intermediate copy number (wcICN) or segmental ICN (sICN) stratified by 10% increments.

| **Live birth, % (n)** | **ICN (wcICN or sICN)** | **Adjusted OR (95%CI, p value)** | **wcICN** | **Adjusted OR (95%CI, p value)** | **sICN** | **Adjusted OR (95%CI, p value)** |
|---|---|---|---|---|---|---|
| **ICN_02-03** | 54.9% (156/284) | 0.84 (0.66-1.08, 0.1657) | 53.6% (60/112) | 0.84 (0.57-1.24, 0.3862) | 55.8% (96/172) | 0.84 (0.61-1.15, 0.2732) |
| **ICN_03-04** | 56.8% (251/442) | 0.89 (0.72-1.08, 0.2388) | 50.8% (92/181) | 0.8 (0.59-1.08, 0.143) | 60.9% (159/261) | 0.95 (0.73-1.24, 0.7276) |
| **ICN_04-05** | 53.7% (180/335) | 0.85 (0.68-1.07, 0.1649) | 48.4% (61/126) | 0.77 (0.53-1.11, 0.1562) | 56.9% (119/209) | 0.9 (0.68-1.21, 0.495) |
| **ICN_05-06** | 44.6% (79/177) | 0.55 (0.4-0.75, 0.0002) | 41.6% (32/77) | 0.77 (0.53-1.11, 0.1562) | 47.0% (47/100) | 0.63 (0.42-0.95, 0.0264) |
| **ICN_>06** | 47.3% (78/165) | 0.62 (0.45-0.85, 0.0032) | 37.7% (20/53) | 0.46 (0.32-0.67, <0.001) | 51.8% (58/112) | 0.71 (0.48-1.04, 0.0808) |

**Supplementary Table 5: LBR per chromosome from the primary US cohort.**

| chr | type | LB = 0 | LB = 1 |
|---|---|---|---|
| chr1 | ICN | 49.41% (42/85) | 50.59% (43/85) |
| | wcICN | 54.55% (12/22) | 45.45% (10/22) |
| | sICN | 47.62% (30/63) | 52.38% (33/63) |
| chr2 | ICN | 38.67% (29/75) | 61.33% (46/75) |
| | wcICN | 46.67% (7/15) | 53.33% (8/15) |
| | sICN | 36.67% (22/60) | 63.33% (38/60) |
| chr3 | ICN | 46.88% (30/64) | 51.56% (33/64) |
| | wcICN | 47.06% (8/17) | 52.94% (9/17) |
| | sICN | 46.81% (22/47) | 51.06% (24/47) |
| chr4 | ICN | 42.5% (34/80) | 57.5% (46/80) |
| | wcICN | 56.52% (13/23) | 43.48% (10/23) |
| | sICN | 36.84% (21/57) | 63.16% (36/57) |
| chr5 | ICN | 49.18% (30/61) | 49.18% (30/61) |
| | wcICN | 50.0% (7/14) | 50.0% (7/14) |
| | sICN | 48.94% (23/47) | 48.94% (23/47) |
| chr6 | ICN | 37.23% (35/94) | 61.7% (58/94) |
| | wcICN | 59.09% (13/22) | 40.91% (9/22) |
| | sICN | 30.56% (22/72) | 68.06% (49/72) |
| chr7 | ICN | 55.68% (49/88) | 43.18% (38/88) |

| | | | |
|---|---|---|---|
| | **wcICN** | 56.67% (17/30) | 43.33% (13/30) |
| | **sICN** | 55.17% (32/58) | 43.1% (25/58) |
| **chr8** | **ICN** | 47.06% (32/68) | 50.0% (34/68) |
| | **wcICN** | 62.5% (15/24) | 37.5% (9/24) |
| | **sICN** | 38.64% (17/44) | 56.82% (25/44) |
| **chr9** | **ICN** | 43.21% (35/81) | 55.56% (45/81) |
| | **wcICN** | 64.71% (11/17) | 35.29% (6/17) |
| | **sICN** | 37.5% (24/64) | 60.94% (39/64) |
| **chr10** | **ICN** | 42.59% (23/54) | 57.41% (31/54) |
| | **wcICN** | 60.0% (9/15) | 40.0% (6/15) |
| | **sICN** | 35.9% (14/39) | 64.1% (25/39) |
| **chr11** | **ICN** | 44.29% (31/70) | 51.43% (36/70) |
| | **wcICN** | 33.33% (3/9) | 66.67% (6/9) |
| | **sICN** | 45.9% (28/61) | 49.18% (30/61) |
| **chr12** | **ICN** | 38.64% (17/44) | 59.09% (26/44) |
| | **wcICN** | 37.5% (6/16) | 62.5% (10/16) |
| | **sICN** | 39.29% (11/28) | 57.14% (16/28) |
| **chr13** | **ICN** | 30.95% (13/42) | 69.05% (29/42) |
| | **wcICN** | 16.67% (3/18) | 83.33% (15/18) |
| | **sICN** | 41.67% (10/24) | 58.33% (14/24) |

| | | | |
|---|---|---|---|
| chr14 | ICN | 46.48% (33/71) | 52.11% (37/71) |
| | wcICN | 50.0% (20/40) | 50.0% (20/40) |
| | sICN | 41.94% (13/31) | 54.84% (17/31) |
| chr15 | ICN | 40.0% (12/30) | 60.0% (18/30) |
| | wcICN | 33.33% (6/18) | 66.67% (12/18) |
| | sICN | 50.0% (6/12) | 50.0% (6/12) |
| chr16 | ICN | 29.63% (8/27) | 70.37% (19/27) |
| | wcICN | 31.25% (5/16) | 68.75% (11/16) |
| | sICN | 27.27% (3/11) | 72.73% (8/11) |
| chr17 | ICN | 55.56% (20/36) | 44.44% (16/36) |
| | wcICN | 72.73% (8/11) | 27.27% (3/11) |
| | sICN | 48.0% (12/25) | 52.0% (13/25) |
| chr18 | ICN | 50.0% (19/38) | 50.0% (19/38) |
| | wcICN | 50.0% (11/22) | 50.0% (11/22) |
| | sICN | 50.0% (8/16) | 50.0% (8/16) |
| chr19 | ICN | 55.0% (11/20) | 45.0% (9/20) |
| | wcICN | 50.0% (7/14) | 50.0% (7/14) |
| | sICN | 66.67% (4/6) | 33.33% (2/6) |
| chr20 | ICN | 53.57% (15/28) | 46.43% (13/28) |
| | wcICN | 53.33% (8/15) | 46.67% (7/15) |

| | | | |
|---|---|---|---|
| | sICN | 53.85% (7/13) | 46.15% (6/13) |
| chr21 | ICN | 47.06% (16/34) | 52.94% (18/34) |
| | wcICN | 42.86% (12/28) | 57.14% (16/28) |
| | sICN | 66.67% (4/6) | 33.33% (2/6) |
| chr22 | ICN | 75.0% (15/20) | 25.0% (5/20) |
| | wcICN | 73.68% (14/19) | 26.32% (5/19) |
| | sICN | 100.0% (1/1) | 0.0% (0/1) |

**Supplementary Table 6:** sex chromosome ICNs and live-birth outcome.

| US.LB | | % (n) | 95%CI |
|---|---|---|---|
| **Euploid** | | 60.3% (4969/8236) | 59.3-61.4% |
| **Mos.Sex** | | 45.7% (42/92) | 35.2-56.4% |
| | X.mos | 40.0% (12/30) | 22.7-59.4% |
| | X.segm | 41.7% (15/36) | 25.5-59.2% |
| | Y.mos | 68.2% (15/22) | 45.1-86.1% |
| **Total** | | 60.2% (5011/8328) | 59.1-61.2% |

| EU.LB | | % (n) | 95%CI |
|---|---|---|---|
| **Euploid** | | 46.3% (2125/4593) | 44.8-47.7% |
| **Mos.Sex** | | 41.3% (19/46) | 27.0-56.8% |
| | X.mos | 45.5% (10/22) | 24.4-67.8% |
| | X.segm | 54.5% (6/11) | 23.4-83.2% |
| | Y.mos | 23.1% (3/13) | 5.0-53.8% |
| **Total** | | 46.2% (2144/4639) | 44.8-47.7% |

**Supplementary Table 7: Pregnancy, obstetrical and neonatal outcomes following the transfer of embryos negative for aneuploidy in the validation European cohort.**

| Pregnancy outcomes per transfer | no ICN (euploid) | ICN (wcICN or sICN) | Adjusted OR (95%CI, p value) | wcICN | Adjusted OR (95%CI, p value) | sICN | Adjusted OR (95%CI, p value) |
|---|---|---|---|---|---|---|---|
| Positive pregnancy test, % (n) | 65.5% (2994/4639) | 59.5% (415/697) | 0.82 (0.69-0.97, 0.0223) | 60.7% (229/377) | 0.88 (0.7-1.1, 0.2521) | 58.1% (186/320) | 0.76 (0.6-0.96, 0.0241) |
| Biochemical pregnancy loss, % (n) | 9.9% (459/4639) | 11.0% (77/697) | 1.11 (0.84-1.46, 0.4667) | 11.1% (42/377) | 1.05 (0.72-1.51, 0.8159) | 10.9% (35/320) | 1.18 (0.81-1.73, 0.3874) |
| Clinical pregnancy, % (n) | 54.6% (2535/4639) | 48.5% (338/697) | 0.8 (0.67-0.94, 0.0079) | 49.6% (187/377) | 0.87 (0.69-1.08, 0.2083) | 47.2% (151/320) | 0.72 (0.57-0.92, 0.0072) |
| Miscarriage, % (n) | 8.4% (391/4639) | 9.6% (67/697) | 1.37 (1.01-1.86, 0.0539) | 10.1% (38/377) | 1.39 (0.94-2.07, 0.1018) | 9.1% (29/320) | 1.35 (0.86-2.1, 0.1909) |
| Ongoing/Live birth, % (n) | 46.2% (2144/4639) | 38.8% (271/697) | 0.75 (0.63-0.89, 0.001) | 39.5% (149/377) | 0.8 (0.64-1.0, 0.0549) | 38.1% (122/320) | 0.69 (0.54-0.89, 0.0035) |
| **Obstetrical and neonatal outcomes per followed-up pregnancies** | | | | | | | |
| Gestational hypertensive disorders, % (n) | 0.6% (12/2117) | 0.0% (0/267) | - | 0.0% (0/147) | - | 0.0% (0/120) | - |
| Preterm delivery, % (n) | 0.9% (19/2117) | 0.0% (0/267) | - | 0.0% (0/147) | - | 0.0% (0/120) | - |
| Low birth weight, % (n) | 4.2% (88/2117) | 4.5% (12/267) | 1.12 (0.6-2.08, 0.728) | 4.1% (6/147) | 1.01 (0.43-2.38, 0.97) | 5.0% (6/120) | 1.25 (0.53-2.94, 0.61) |
| NICU admission, % (n) | 3.1% (66/2117) | 2.6% (7/267) | 0.89 (0.4-1.96, 0.7657) | 1.4% (2/147) | 0.48 (0.12-1.99, 0.31) | 4.2% (5/120) | 1.54 (0.60-3.94, 0.37) |
| Congenital anomalies, % (n) | 0.8% (16/2117) | 0.4% (1/267) | 0.61 (0.08-4.69, 0.6367) | 0.0% (0/147) | - | 0.8% (1/120) | 1.38 (0.18-10.79, 0.76) |
