## supplementary for "PGT-A Mosaicism Reporting Lacks Clinical Predictive Value: Evidence from a Multisite, Double-Blinded Study with Independent Validation Across 15,315 Single-Embryo Transfers"

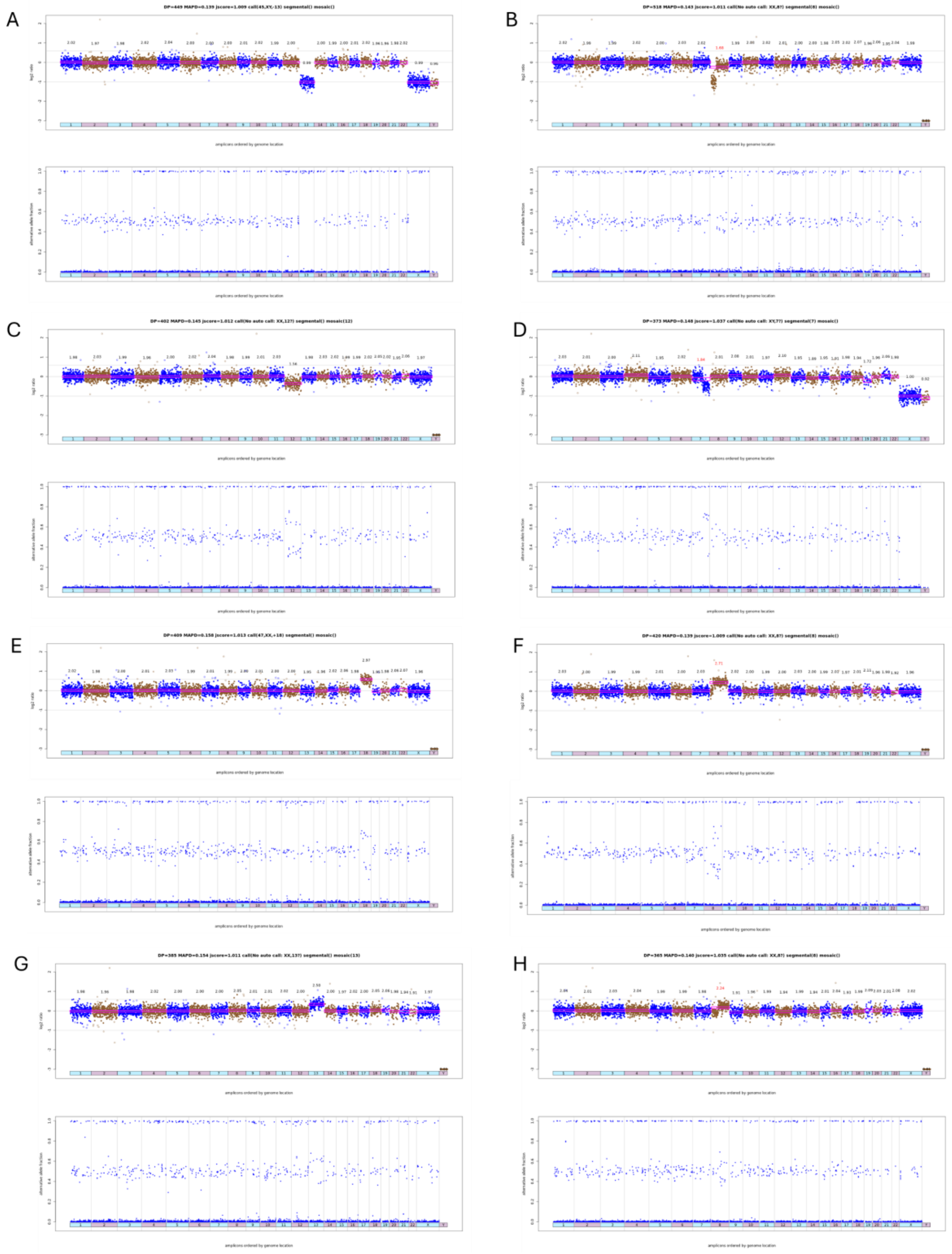
A
DP=449 MAPD=0.139 jscore=1.009 call(45,XY,-13) segmental() mosaic()
B
DP=518 MAPD=0.143 jscore=1.011 call(No auto call: XX,8?) segmental(8) mosaic()
C
DP=402 MAPD=0.145 jscore=1.012 call(No auto call: XX,12?) segmental() mosaic(12)
D
DP=373 MAPD=0.148 jscore=1.037 call(No auto call: XY,7?) segmental(7) mosaic()
E
DP=409 MAPD=0.158 jscore=1.013 call(47,XX,+18) segmental() mosaic()
F
DP=420 MAPD=0.139 jscore=1.009 call(No auto call: XX,8?) segmental(8) mosaic()
G
DP=385 MAPD=0.154 jscore=1.011 call(No auto call: XX,13?) segmental() mosaic(13)
H
DP=365 MAPD=0.140 jscore=1.035 call(No auto call: XX,8?) segmental(8) mosaic()
amplicons ordered by genome location

**Supplementary Figure 2:** (A) aneuploidy and ICN patterns per female age in primary U.S. cohort, (B) aneuploidy and ICN patterns per female age in validation European cohort, (C) aneuploidy patterns per chromosome in primary U.S. cohort.

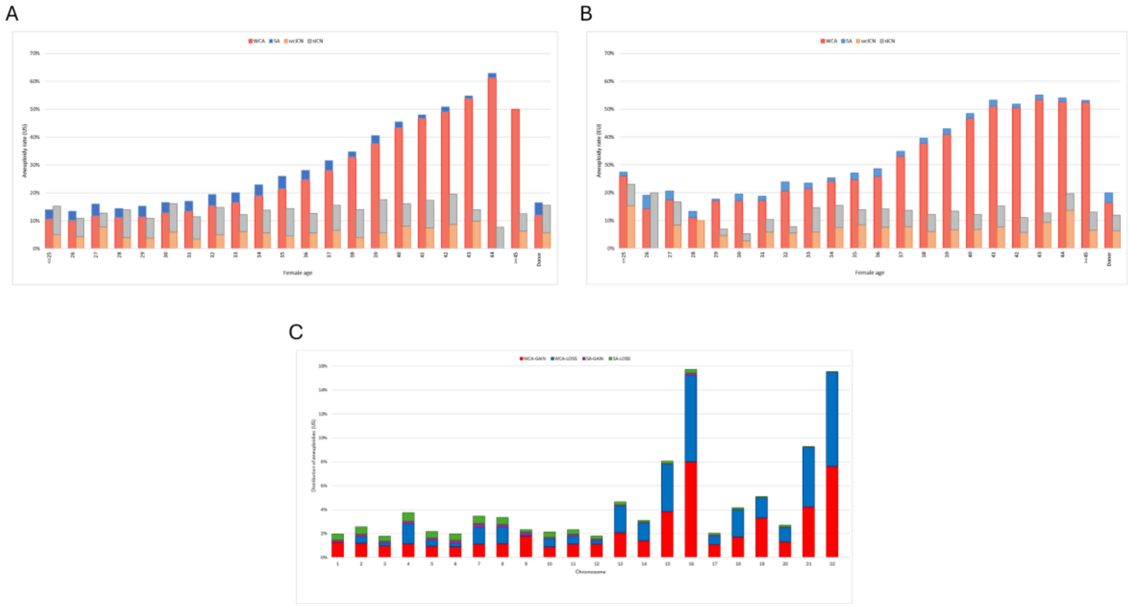


**Supplementary Figure 3.** Distribution of (A) wcICNs and (B) sICNs per chromosome in primary U.S. cohort after unblinding and their correlation with MAPD (C).

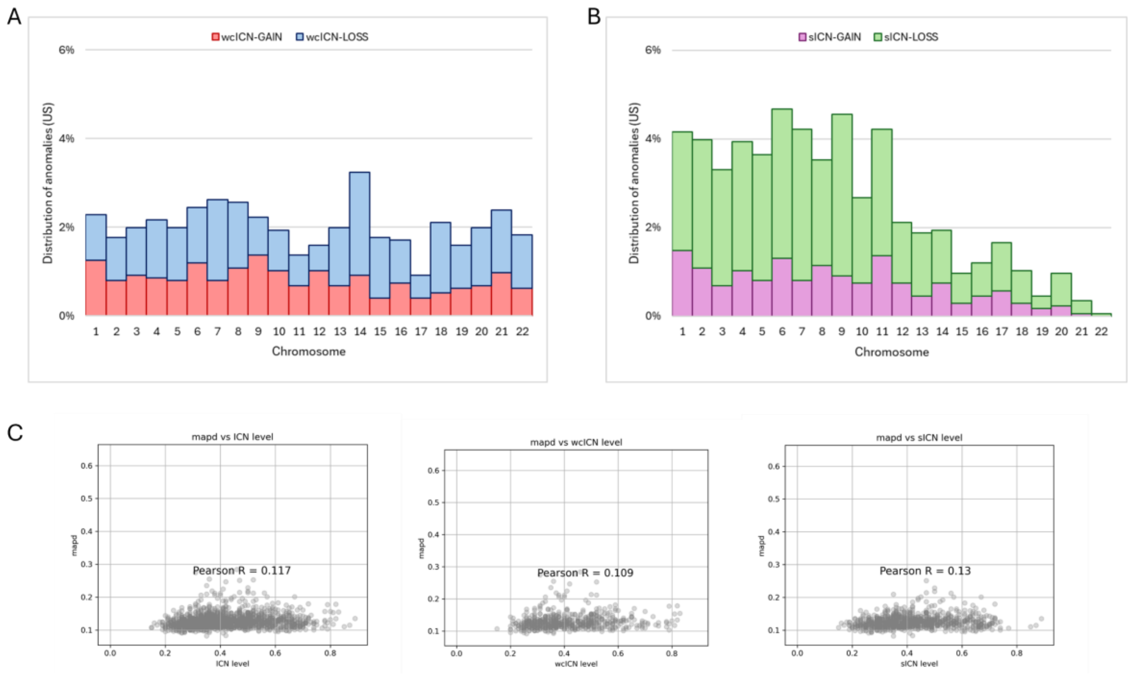

**Supplementary Figure 4: Association of ICN with LBR in primary U.S. cohort analysis** according to low and high grade with a threshold of 50% as maximum ICN deviation from the standard disomy state (A). Outcomes are further stratified by 10% increments in intermediate copy number (ICN) for both segmental (B) and whole-chromosome (C) variations.

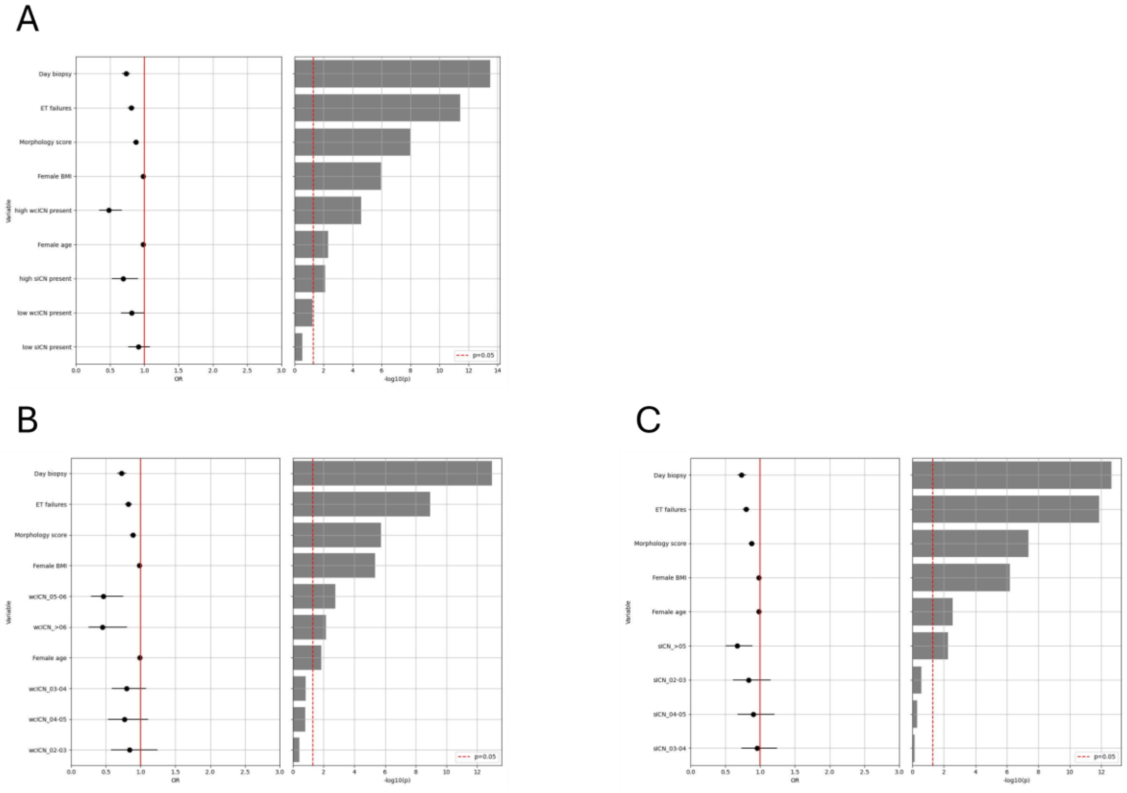

**Supplementary Figure 5: Impact of ICN reporting on LBR in primary U.S. cohort; analysis based on ROC analysis,** for (A) wcICN; (B) sICN; (C) high- wcICN only, (D) high-sICN only, (E) high wcICN and high-sICN combined.

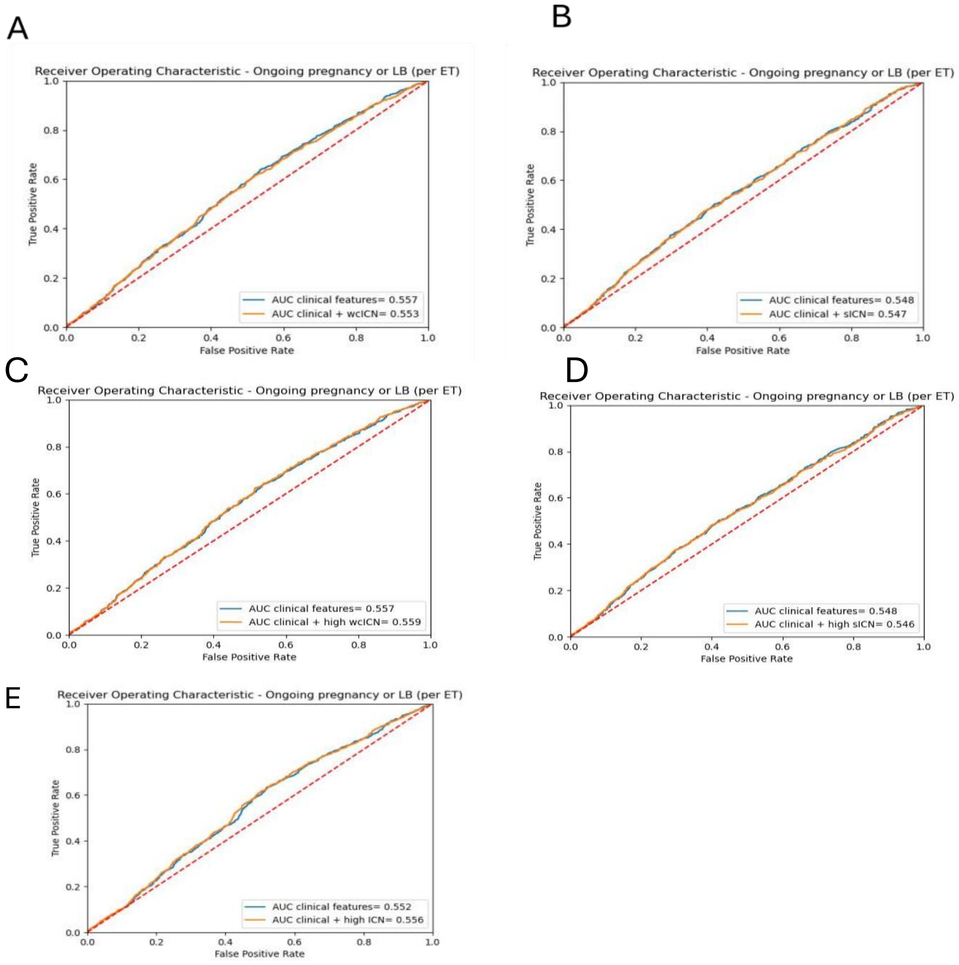

**Supplementary Figure 6. Association of ICN with LBR in the validation European cohort analysis.** (A) Logistic regression analysis including covariates. (B) ROC curve. (C) Decision tree.

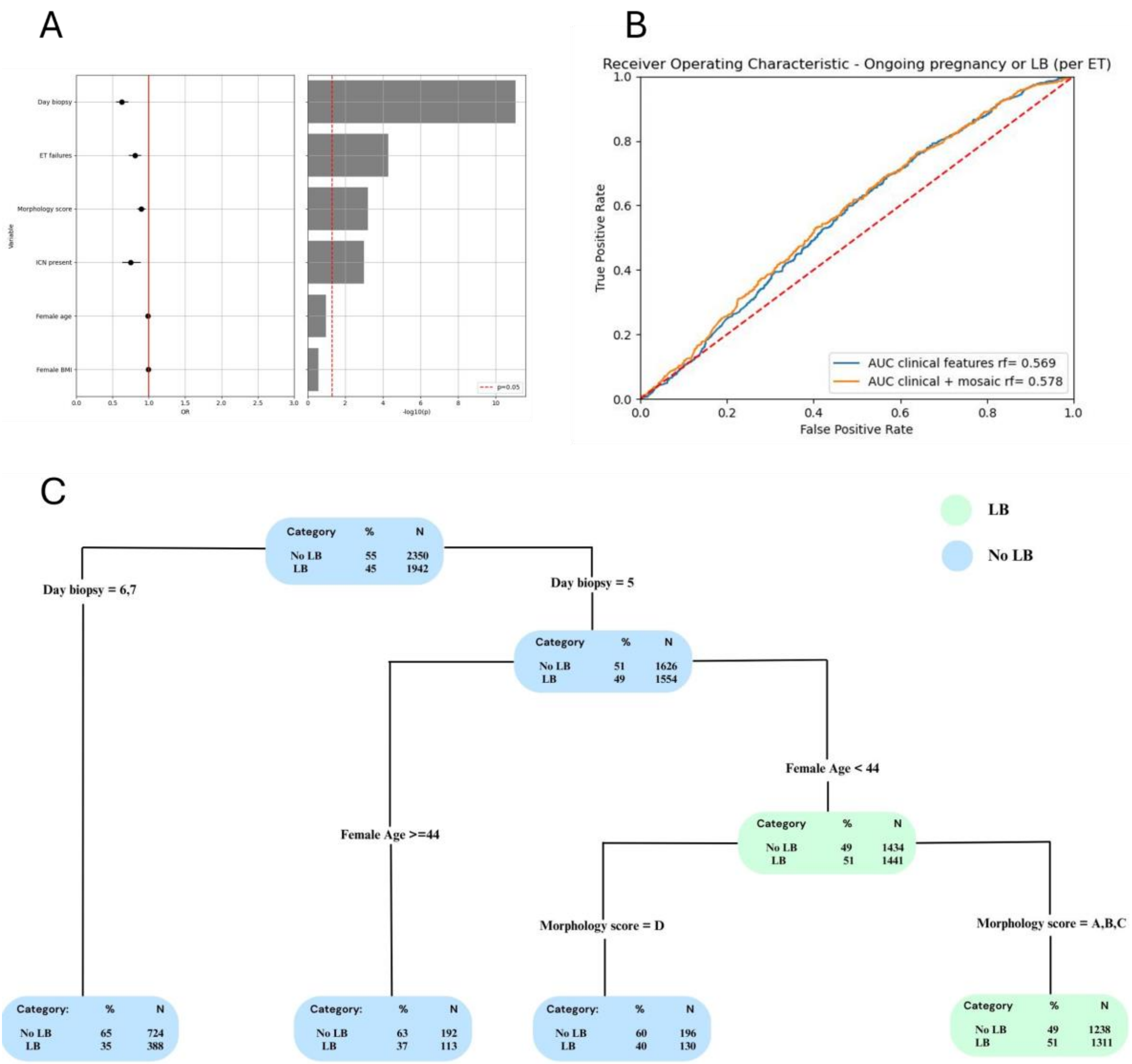

**Supplementary Table 1. ICN validation**. Experimental mixtures of cell lines (GM01359 carrying 47,XY,+18 and GM02948 carrying 47,XY,+13) at different proportions to mimic chromosomal mosaicism of different levels. CN=copy number value, SD=standard deviation, wcICN-GAIN=intermediate copy number of a whole chromosome towards trisomy.
